# Treatment with Soluble CD24 Attenuates COVID-19-Associated Systemic Immunopathology

**DOI:** 10.1101/2021.08.18.21262258

**Authors:** No-Joon Song, Carter Allen, Anna E. Vilgelm, Brian P. Riesenberg, Kevin P. Weller, Kelsi Reynolds, Karthik B. Chakravarthy, Amrendra Kumar, Aastha Khatiwada, Zequn Sun, Anjun Ma, Yuzhou Chang, Mohamed Yusuf, Anqi Li, Cong Zeng, John P. Evans, Donna Bucci, Manuja Gunasena, Menglin Xu, Namal P.M. Liyanage, Chelsea Bolyard, Maria Velegraki, Shan-Lu Liu, Qin Ma, Martin Devenport, Yang Liu, Pan Zheng, Carlos D. Malvestutto, Dongjun Chung, Zihai Li

**Affiliations:** The Ohio State University, Columbus, OH, USA; Department of Internal Medicine, The Ohio State University College of Medicine, Columbus, OH; The Pelotonia Institute for Immuno-Oncology, The Ohio State University Comprehensive Cancer Center, Columbus, OH, USA; Dept of Biomedical Informatics, The Ohio State University College of Medicine, Columbus, OH; Department of Public Health Sciences, Medical University of South Carolina, Charleston, SC; The Ohio State University Comprehensive Cancer Center, Columbus, OH; Department of Microbiology, The Ohio State University College of Arts and Sciences, Columbus, OH, USA; Department of Microbial Infection and Immunity, The Ohio State University College of Medicine, Columbus, OH, USA; Department of Veterinary Biosciences, The Ohio State University College of Veterinary Medicine, Columbus, OH, USA; Department of Pathology, The Ohio State University College of Medicine, Columbus, OH; The Ohio State University College of Medicine, Columbus, OH, USA; Center for Retrovirus Research and Department of Veterinary Biosciences, The Ohio State University, Columbus, OH, USA; OncoC4, Rockwille, MD, USA

## Abstract

**BACKGROUND:** SARS-CoV-2 causes COVID-19 through direct lysis of infected lung epithelial cells, which releases damage-associated molecular patterns (DAMPs) and induces a pro-inflammatory cytokine milieu causing systemic inflammation. Anti-viral and anti-inflammatory agents have shown limited therapeutic efficacy. Soluble CD24 (CD24Fc) is able to blunt the broad inflammatory response induced by DAMPs in multiple models. A recent randomized phase III trial evaluating the impact of CD24Fc in patients with severe COVID-19 demonstrated encouraging clinical efficacy.

**METHODS:** We studied peripheral blood samples obtained from patients enrolled at a single institution in the SAC-COVID trial (NCT04317040) collected before and after treatment with CD24Fc or placebo. We performed high dimensional spectral flow cytometry analysis of peripheral blood mononuclear cells and measured the levels of a broad array of cytokines and chemokines. A systems analytical approach was used to discern the impact of CD24Fc treatment on immune homeostasis in patients with COVID-19.

**FINDINGS:** Twenty-two patients were enrolled, and the clinical characteristics from the CD24Fc vs. placebo groups were matched. Using high-content spectral flow cytometry and network-level analysis, we found systemic hyper-activation of multiple cellular compartments in the placebo group, including CD8^+^ T cells, CD4^+^ T cells, and CD56^+^ NK cells. By contrast, CD24Fc-treated patients demonstrated blunted systemic inflammation, with a return to homeostasis in both NK and T cells within days without compromising the ability of patients to mount an effective anti-Spike protein antibody response. A single dose of CD24Fc significantly attenuated induction of the systemic cytokine response, including expression of IL-10 and IL-15, and diminished the coexpression and network connectivity among extensive circulating inflammatory cytokines, the parameters associated with COVID-19 disease severity.

**INTERPRETATION:** Our data demonstrates that CD24Fc treatment rapidly down-modulates systemic inflammation and restores immune homeostasis in SARS-CoV-2-infected individuals, supporting further development of CD24Fc as a novel therapeutic against severe COVID-19.

**FUNDING:** NIH

## INTRODUCTION

The pathogenesis of SARS-CoV-2 is a multistep process starting with the infection of ACE2-expressing lung epithelial cells^1^. Following infection, unconstrained viral replication leads to cell lysis and the release of DAMPs. Recognition of these molecules by neighboring cells produces a pro-inflammatory milieu through the release of cytokines (such as IL-6 and IL-10), which recruit and activate monocytes, macrophages, and T cells^2^. In severe COVID-19, this pro-inflammatory feedback loop results in a persistent and harmful response that leads to structural damage of the lung. The resulting cytokine storm can lead to acute respiratory distress syndrome (ARDS), multi-organ failure and death^3^.

Even though COVID-19 mRNA vaccines have shown great success in preventing severe disease^4^, recent reports suggest that new SARS-CoV-2 variant delta can escape from the immune response induced by existing vaccines^5^. Breakthrough infections post full vaccinations can occur^6^, especially in immunocompromized individuals^7^, requiring urgent development of effective therapeutic agents against this disease. Interim results from the Solidarity trial (NCT04315948) indicate that several repurposed interventions do not significantly alter COVID-19 morbidity and mortality^8^. Other approaches, including cytokines and convalescent plasma, have also been largely ineffective^9, 10^. The anti-inflammatory glucocorticoid dexamethasone is one of the few interventions shown to reduce mortality in patients with critical-to-severe COVID-19^11^.

CD24Fc treatment attenuates inflammation associated with viral infections, autoimmunity, and graft-versus-host diseases^12–14^. In this study, we compared blood samples from COVID-19 patients enrolled in the SAC-COVID trial following CD24Fc or placebo. We examined dynamic changes in peripheral blood mononuclear cells (PBMCs) and systemic cytokine and chemokine levels. We demonstrated that CD24Fc reversed the inflammatory hallmarks associated with severe COVID-19, including cytokine storm and immune hyperactivation.

## METHODS

### PATIENTS AND TRIAL PROCEDURE

This study included samples from patients enrolled in NCT04317040 at The Ohio State University Wexner Medical Center (patient details described in Table S1). Patients eligible for this trial were hospitalized with COVID-19, requiring supplemental oxygen but not mechanical ventilation, with a prior positive SARS-CoV-2 PCR test. Consented and enrolled patients were randomized in a double-blinded fashion to receive either CD24Fc antibody (480 mg IV infusion) or placebo control (IV saline). Peripheral blood samples were collected from patients before (day 1, D1) and after (D2, D4, D8, D15, and D29) treatment. The Western Institutional Review Board approved trial and protocol. The study was monitored by a contract research organization; safety reports were submitted to an independent Data and Safety Monitoring Board. This trial was conducted in compliance with the protocol, International Conference on Harmonization Good Clinical Practice, and all applicable regulatory requirements.

### LABORATORY ASSAYS

Immune profiling, viral neutralization, and cytokine/chemokine assays were performed at The Ohio State University, and per manufacturer’s instructions as applicable^15, 16^. We developed multiple high dimensional spectral flow cytometry panels to study the dynamic changes of CD8^+^, CD4^+^, and CD56^+^ immune cell subsets (Table S2). See Supplementary Appendix for details.

### BIOINFORMATICS AND STATISTICAL ANALYSIS

Bioinformatic analyses were performed as previously described^17–25^. Flow cytometry data were preprocessed using the OMIQ software, visualized using the Uniform Manifold Approximation and Projection (UMAP) algorithm, and analyzed using a multivariate t-mixture model^17^. Immune cell activation score was constructed by aggregating pre-selected activation markers^18, 19^ using a principal component analysis (PCA) applied to the flow cytometry data of

HD and baseline COVID-19 patients. Cytokine score was constructed using a weighted sum approach and validated using PCA and autoencoder approaches^20^. Network-level analysis of cytokine data was implemented by constructing a correlation network between cytokines and evaluating the network structure and importance of each node in the network based on an eigenvector centrality (EC) score^24^. Group comparisons were evaluated using independent sample t-test or Kruskal-Wallis test for continuous variables, and Chi-squared test for categorical variables. Longitudinal analyses were implemented using generalized linear mixed models (GLMMs).

## RESULTS

### POPULATION DYNAMICS OF IMMUNE CELLS

e utilized a high dimensional spectral flow cytometry panel with an extensive array of immune population markers (Table S2) to analyze the systemic effects of SARS-CoV-2 and CD24Fc treatment on PBMCs. Using an unbiased clustering approach based on a multivariate *t*-mixture model^17^, we identified 12 distinct clusters that we visualized in two dimensions using the UMAP algorithm (Fig 1A). Using clustered heatmap analysis, we correlated expression intensity with clusters to annotate B cells (clusters 1, 6, 8), CD8^+^ T cells (clusters 7, 11, 12), CD4^+^ T cells (clusters 2, 3), γδ T cells (cluster 4), NK cells (cluster 10), and myeloid cells (clusters 5, 9) (Fig 1B). Comparing systemic immune population dynamics (Fig 1C-D), we found significant increases in plasma B cells (cluster 6), NK cells (cluster 10), and terminally differentiated CD8^+^ T cells (cluster 12) in baseline (D1) COVID-19 patients vs. healthy donors (HD). Conversely, we found that HD samples were enriched for naïve CD8^+^ T cells (cluster 11) and a subset of myeloid cells (cluster 5). These initial findings were consistent with established immunopathology of SARS-CoV-2 infection and the important role the adaptive immune system plays in viral pathogen response^26–29^, and thus validated our experimental approach.

**Figure 1.**
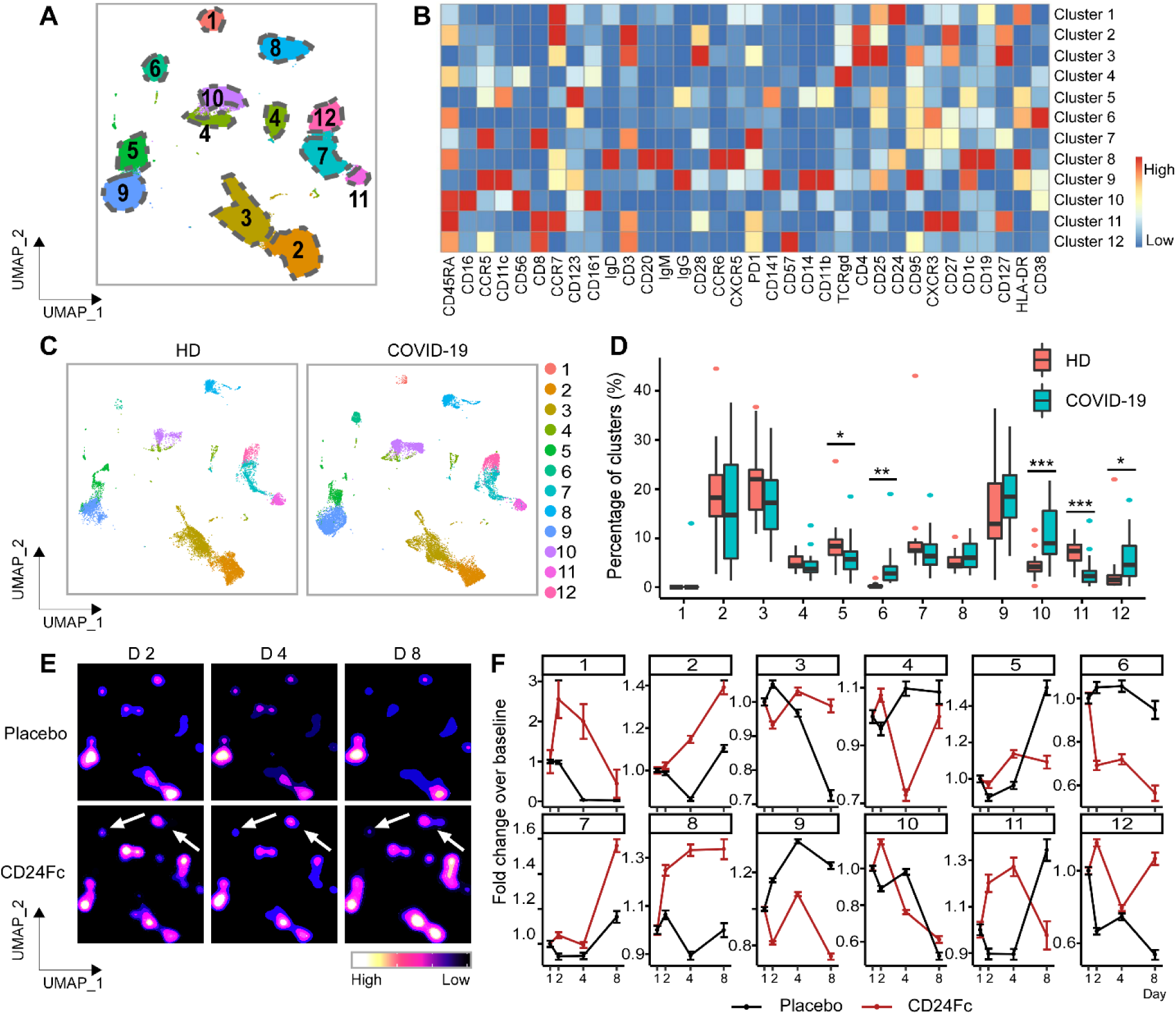
Population dynamics of peripheral blood mononuclear cells from healthy donors vs. patients with COVID-19 treated with placebo or CD24Fc. A total of 1,306,473 PBMCs from HD (n=17) and COVID-19 patients (n=22) were clustered using an unbiased multivariate *t*-mixture model, which identified 12 sub-clusters that reflect statistically distinct cell states. Visualization of the relative similarity of each cell and cell cluster on the two-dimensional UMAP space with a 10% downsampling (**Panel A**). Cluster-by-marker heatmap characterizing the expression patterns of individual clusters (**Panel B**). UMAP dot plots (**Panel C**) and cluster frequencies (**Panel D**) of HD vs. baseline COVID-19 patient samples (cluster 5, p=0.03; cluster 6, p=0.001; cluster 10, p<0.001; cluster 11, p<0.001). Contour plots representing the density of cells throughout regions of the UMAP space from COVID-19 patients D2, D4, and D8 after CD24Fc vs. placebo treatment (**Panel E**, white arrows indicate visual changes between CD24Fc vs. placebo contour plots). Selected cluster population dynamics as fold change over baseline for each group over time (**Panel F**) (D2: placebo n=12, CD24Fc n=10; D4: placebo n=11, CD24Fc n=9; D8: placebo n=4, CD24Fc n=3). The p-value was calculated using the Kenward-Roger method. **\***, p<0.05; **\*\***, p<0.01; **\*\*\***, p<0.001.

We next used UMAP contour plots to investigate the effects of CD24Fc treatment on immune population dynamics over time (Fig 1E-F). From baseline to D8, the CD24Fc group displayed a sharp and steady decline of plasma B cells (cluster 6), which coordinated with a proportional increase in mature B cells (cluster 8). The placebo group showed relatively stable cell proportions for these populations over the same time frame. There were no significant differences between the two groups in mounting an effective anti-Spike protein antibody response (Fig S1).

### CD24Fc SUPPRESSES T CELL ACTIVATION

We developed a 25-marker flow cytometry panel to examine the intricacies associated with effector cell (NK and CD4**^+^**/CD8**^+^** T cell) activation and differentiation in response to SARS-CoV-2 infection and CD24Fc treatment (Table S2). Using our unbiased clustering approach, we identified eight distinct clusters within CD8**^+^** T cells from COVID-19 and HD samples (Fig 2A-C). At baseline, COVID-19 samples showed enriched frequency of clusters 4, 5, 7, and 8, which express markers of activation; HD samples were skewed towards cluster 1, which exhibits a naive phenotype (Fig. 2D-E). To analyze the impact of CD24Fc on CD8**^+^** T cell activation, we generated UMAP contour plots for each treatment group (Fig 2F), and analyzed changes to cluster proportions over time (Fig 2G). CD24Fc treatment correlated with a modest increase in cluster 1 frequency over time, whereas placebo-treated patients showed marked decline. Conversely, the proportion of cluster 8 cells (a population whose expression pattern is suggestive of highly activated CD8^+^ T cells) were stagnant in CD24Fc-treated patients, compared to the marked increase seen in placebo group (Fig 2G).

**Figure 2.**
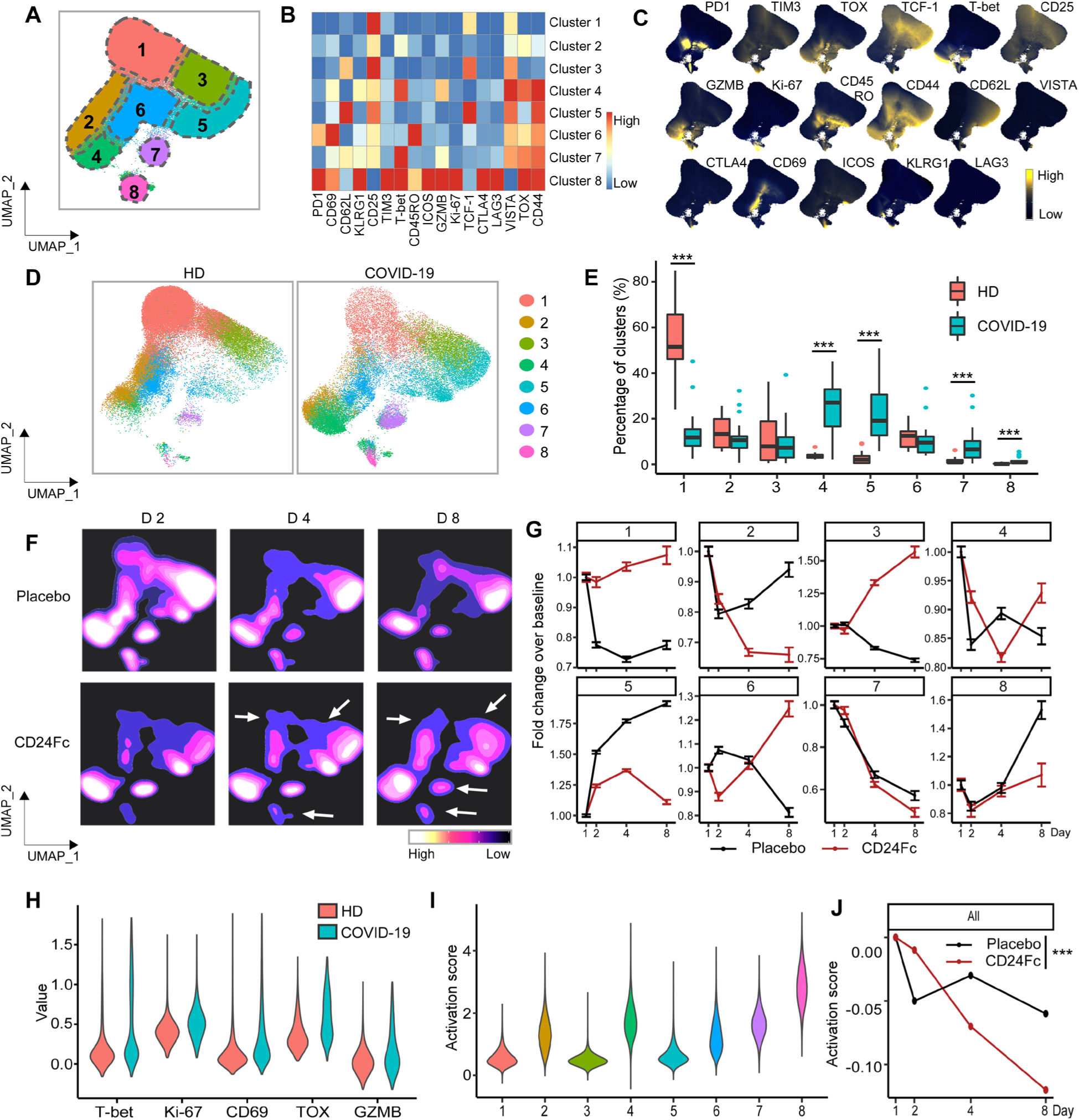
Subcluster analysis of peripheral blood CD8^+^ T cells in COVID-19 patients: activation following SARS-CoV2 infection is dampened by CD24Fc treatment. A total of 1,466,822 CD8^+^ cells from HD (n=17) and COVID-19 (n=22) patients were clustered using an unbiased multivariate *t*-mixture model, which identified 8 CD8^+^ sub-clusters that reflect statistically distinct CD8^+^ T cell activation states. The relative similarity of each cell and cell cluster on the two-dimensional UMAP space were visualized with a 10% downsampling (**Panel A**). Using median expression of flow cytometry markers, a cluster-by-marker heatmap was generated to characterize the subsets (**Panel B**) and visualize individual marker expression patterns on the UMAP space (**Panel C**). To understand the effect of SARS-CoV2 infection on cell population dynamics, a comparison was made with UMAP dot plots (**Panel D**) and cluster frequencies (**Panel E**) of HD vs. baseline COVID-19 patient samples (cluster 1, p<0.001; cluster 4, p<0.001; cluster 5, p<0.001; cluster 7, p<0.001; cluster 8, p<0.001). The samples from COVID-19 patients 2, 4, and 8 days after CD24Fc vs. placebo treatment were displayed using contour plots to represent the density of cells throughout regions of the UMAP space (**Panel F**, white arrows indicate visual changes between CD24Fc vs. placebo contour plots). The cluster population dynamics as fold change over baseline in each treatment group was shown (**Panel G**; sample distribution described in Fig 1F legend). To better characterize the activation status of CD8 T cells, a subset of markers (T-bet, Ki-67, CD69, TOX, GZMB) was linearly transformed to create a univariate cell-level activation score (**Panel H**), where highly activated cell clusters (such as cluster 8) had highest activation scores (**Panel I**). A GLMM was then to fit to the longitudinal cell-level activation scores to assess the effect of CD24Fc treatment on activation scores over time (**Panel J**). The p-value was calculated using the Kenward-Roger method. **\*\*\***, p<0.001.

While tracking cluster proportions over time provides an unbiased global view of the data, these statistically-distinct cell clusters may not always correspond perfectly to biologically-distinct cell types. Therefore, we augmented the unbiased clustering analysis with a semi-supervised approach to define an unbiased CD8^+^ T cell activation score. Known markers of T cell activation (T-bet, Ki-67, CD69, TOX, and GZMB) were significantly increased in baseline COVID-19 patients compared to HD (Fig 2H), supporting our hypothesis that SARS-CoV-2 infection increases peripheral T cell activation. To create a unified cell-

level activation score, we used PCA to implement dimension reduction of the cell-by-activation marker expression data for all baseline COVID-19 and HD cells. The first principal component (PC1) loadings of each activation marker were used as coefficients in a linear model for defining the activation score (Table S3). Thus, while we manually selected key T cell activation markers, we determined the relative contribution of each activation marker to the final activation score in an unbiased and data-adaptive manner, yielding a semi-supervised approach. We observed positive PC1 loadings and positive average log-fold changes for each activation marker, confirming that higher activation scores reflect higher T cell activation (Table S3). Distributions of activation scores across cell clusters also confirmed that more highly activated cell subsets feature higher activation scores (Fig 2I).

To characterize the effect of CD24Fc treatment on global CD8**^+^** T cell activation, we adopted a GLMM of the activation scores over time. While CD8**^+^** T cell activation scores at baseline were not statistically different between groups, the predicted mean activation scores indicate significantly different trajectories between placebo and CD24Fc groups over time (Fig 2J). Thus, we conclude that CD24Fc treatment significantly reduced CD8^+^ T cell activation compared to placebo. CD4^+^ T cell activation also plays an important role in immune response to SARS-CoV-2 infection, so we applied the analysis strategy presented above to this population^26^. To comprehensively understand the role of CD4^+^ T cells and FOXP3^+^ Tregs, we analyzed total CD4^+^ T cells including FOXP3^+^ subset (Fig S2), and then FOXP3^+^ Tregs exclusively (Fig S3). Both analyses showed hyperactivated subsets and overall activation score decreased by CD24Fc treatment.

### CD24Fc REDUCES NK CELL ACTIVATION

The increased number of NK cells in samples from patients with COVID-19 (Fig 1C-D, cluster 10) implies they play an important role in SARS-CoV-2 infection. We investigated the activation status of NK cells using our unbiased clustering and visualization approach, and identified 12 statistically-distinct NK cell clusters, which we visualized on heatmaps and UMAPs (Fig 3A-C). Cluster 5, the most highly represented cluster in HD samples, displayed an expression pattern suggestive of a less activated population. Samples from COVID-19 patients revealed significant reduction in cluster 5 and expansion of clusters 4, 6, 8, 9, 10, 11, and 12 (Fig 3D-E).

**Figure 3.**
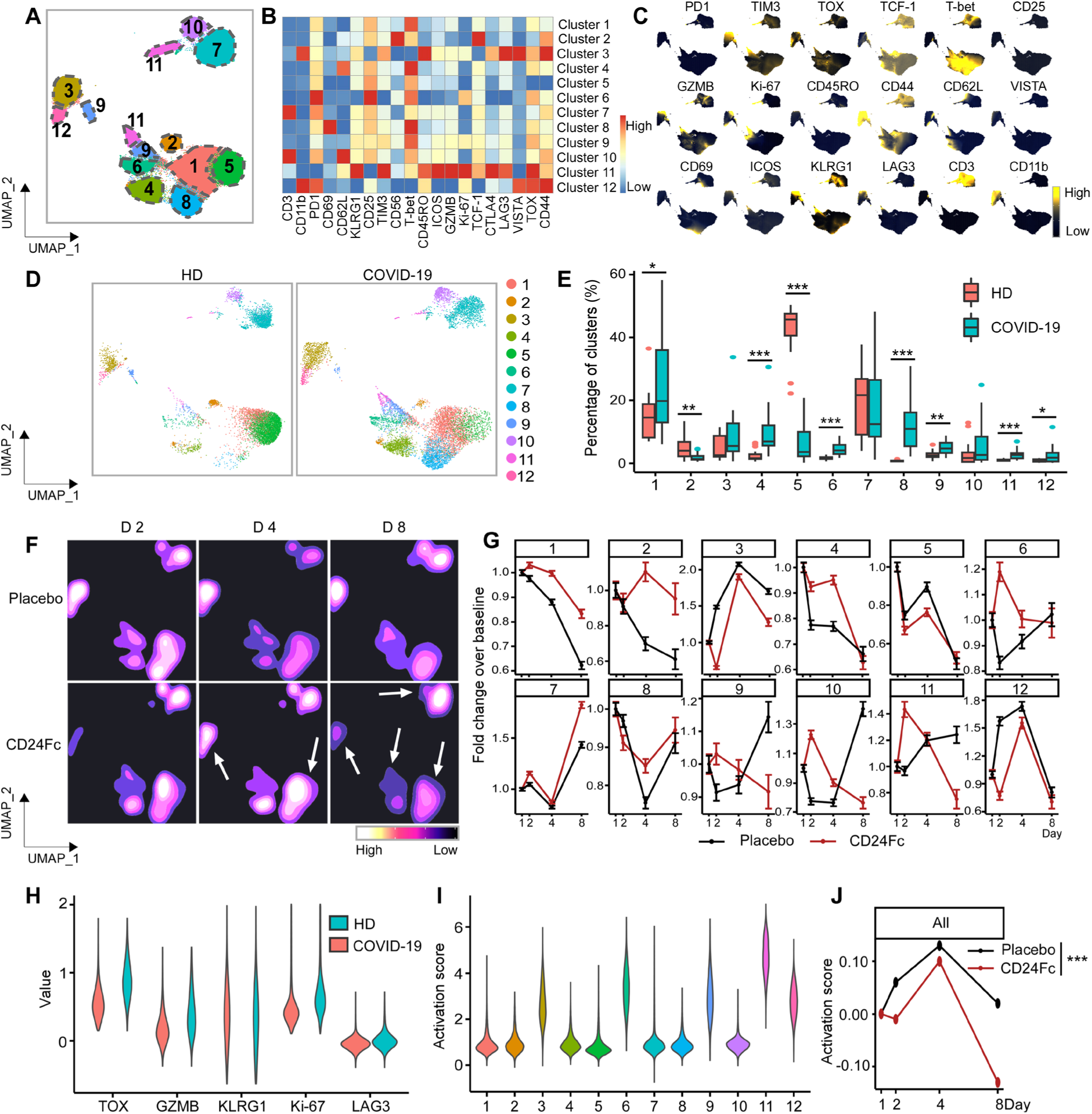
Subcluster analysis of peripheral blood NK cells in COVID-19 patients: activation of following SARS-CoV2 infection is dampened by CD24Fc treatment. CD56^+^ cells (n=783,623) from HD (n=17) and COVID-19 (n=22) patients were clustered using an unbiased multivariate *t*-mixture model, which identified 12 sub-clusters that reflect statistically distinct CD56^+^ T cell activation states. The relative similarity of each cell and cell cluster on the two-dimensional UMAP space were visualized with a 10% downsampling (**Panel A**). Using median expression of flow cytometry markers, a cluster-by-marker heatmap were generated to characterize the subsets (**Panel B**) and visualize individual marker expression patterns on the UMAP space (**Panel C**). To understand the effect of SARS-CoV2 infection on NK cell population dynamics, a comparison was made with UMAP dot plots (**Panel D**) and cluster frequencies (**Panel E**) of HD vs. baseline COVID-19 patient samples. The day 2, 4, 8 samples from placebo and CD24Fc-treated patient groups were visualized using contour plots to represent the density of cells throughout regions of the UMAP space (**Panel F**, white arrows indicate visual changes between CD24Fc vs. placebo contour plots). The cluster population dynamics as fold change over baseline in each treatment group was shown (**Panel G**; sample distribution described in Fig 1 legend). To better characterize the activation status of NK cells, a subset of markers (TOX, GZMB, KLRG1, Ki-67, LAG-3) was linearly transformed to create a univariate cell-level activation score (**Panel H**), where highly activated cell clusters (such as cluster 11) had highest activation scores (**Panel I**). A GLMM was then fit to the longitudinal cell- level activation scores to assess the effect of CD24Fc treatment on activation scores over time (**Panel J**). The p-value was calculated using the Kenward-Roger method. **\***, p<0.05; **\*\***, p<0.01; **\*\*\***, p<0.001.

To understand the role of CD24Fc treatment on NK cell population dynamics, we generated UMAP contour plots to visualize temporal and treatment-based changes (Fig 3F), and quantified these differences (Fig 3G). Clusters 1 and 2, which show mild activation, were increased by CD24Fc, while cluster 11, which expresses multiple activation markers, was decreased. To visualize activation, known NK cell activation markers (TOX, GZMB, KLRG1, Ki-67, and LAG3) were assessed (Fig 3H) and plotted per cluster (Fig 3I). Using a GLMM of activation scores over time, we found that while baseline values for NK cell activation were not statistically different, the mean activation scores were significantly different between placebo and CD24Fc groups throughout the study duration (Fig 3J). Thus, CD24Fc treatment rapidly reduced NK cell activation status, and the impact was sustained throughout the study period.

### CD24Fc ATTENUATES SYSTEMIC CYTOKINE RESPONSE

To examine the effect of CD24Fc on cytokine response to SARS-CoV-2 infection, we compared plasma cytokine concentrations from HD and COVID-19 patients treated with CD24Fc or placebo. We used multiplex ELISA and Luminex analysis platforms testing 37 cytokines in total. Fifteen out of 37 tested cytokines were significantly elevated during SARS-CoV-2 infection (Fig 4A, Fig S4A). These included cytokines associated with type 1 (IL-12p40, CXCL9, IL-15) and type 3 (IL-1α, IL-1β, RANTES) immunity, and chemokine MCP-1 (CCL2) that recruits monocytes and T cells to the sites of inflammation. Only three out of 37 cytokines were significantly downregulated in COVID-19 patients (Fig S4A).

**Figure 4.**
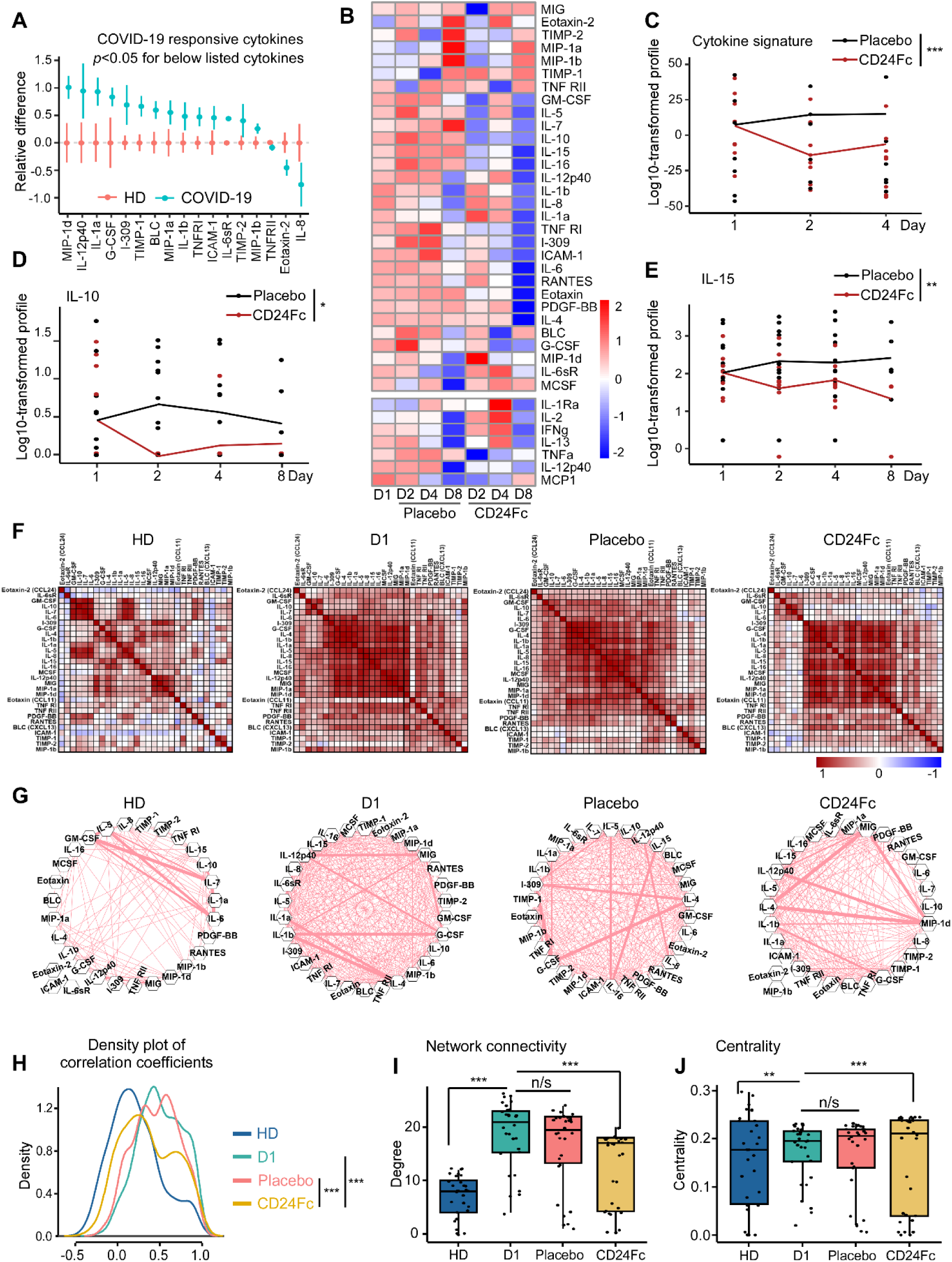
CD24Fc treatment downregulates systemic cytokine response in patients with COVID-19. The relative differences in plasma concentrations of cytokines/chemokines between HD (n=25) and COVID-19 patients (n=22) is shown. Values were log-transformed and evaluated using independent sample t-test. Only significantly up- and down-regulated markers are shown (**Panel A**). The heatmap analysis (**Panel B**) was used to visualize the relative levels of plasma cytokines/chemokines in placebo vs. CD24Fc- treated patients at indicated time points (Placebo: D1 n=12, D2 n=12, D4 n=11, D8 n=5; CD24Fc: D1 n=10, D2 n=10, D4 n=9, D8 n=3). To compare longitudinal patterns across groups, each cytokine had its group-specific baseline mean adjusted to match the overall mean at D1 and consequent time points are normalized accordingly, followed by scaling-by-row. The cytokine score was analyzed longitudinally using weighed sum approach (**Panel C**; p<0.001). Using log-10 transformation of cytokine concentrations (dots) and GLMM predicted fixed effects trends (lines), the changes in IL-10 (**Panel D**; p=0.05) and IL-15 (**Panel E**; p=0.002) levels in CD24Fc (red) and placebo (black) groups were revealed. Values and trend lines were centered at D1 mean. The p-value was calculated using the Kenward-Roger method. Using Pearson correlation matrices (**Panel F**; darker red indicates stronger correlation) and network maps (**Panel G**; weight of edge represents correlation coefficient), 30 plasma markers in HD (n=25), COVID-19 baseline (D1, n=22), placebo (pooled D2-D8, n=28), and CD24Fc-treated (pooled D2-D8, n=24) groups were visualized. Using these correlation coefficients, a density plot between 30 plasma cytokines (**Panel H**; D1 vs placebo, p=0.07; D1 vs CD24Fc, p<0.001; placebo vs CD24Fc, p<0.001) was constructed. Kolmogorov-Smirnov test was used to evaluate equality of densities between groups. Analysis of connectivity (**Panel I**) and centrality analysis of cytokine network (**Panel J**) display the cytokine expression relationships within each group. Network connectivity plots display highly correlated connections for each cytokine (i.e., node degree) and evaluated using paired t-test. Centrality analysis of cytokine network used eigenvector centrality score that considers global network connectivity and correlation coefficients between cytokines (HD vs D1, p<0.001; D1 vs placebo, p=0.08; D1 vs CD24Fc, p<0.001). Bartlett’s test was performed to evaluate the significance of variance of centrality scores (HD vs D1, p=0.013; D1 vs placebo, p=0.17; D1 vs CD24Fc, p=0.008). Each dot in Panel I and J represents a cytokine. *, p<0.05; **, p<0.01; ***, p<0.001.

We next studied the impact of CD24Fc on cytokine expression in patients with COVID-19. As shown in Fig 4B, substantial reduction of cytokines (GM-CSF, IL-5, IL-7, IL-10) and chemokines (MIG, MIP-1α, MIP-1β) was observed within 24 hours of CD24Fc. At 1 week after treatment, most of the cytokines and chemokines tested are reduced by 10-fold or more. The majority of them are selectively reduced in the CD24Fc-treated patients including cytokines critically involved in COVID-19 pathogenesis, such as IL-6 and GM-CSF^30^. Analysis of cytokine scores calculated by integrating expression of all markers tested by multiplex ELISA platform using weighted sum approach demonstrated significant decrease in CD24Fc- treated groups compare to placebo (Fig 4C). This finding was independently confirmed using Autoencoder^20^ and PCA (Fig S4D).

To better understand this modulation, we studied correlations between individual cytokines across groups. Correlation matrices (Fig 4F) showed that only a few groups of cytokines were co-expressed by HD. The numbers of co-regulated cytokines dramatically increased in baseline COVID-19 samples indicating activation of coordinated cytokine response. Remarkably, samples from CD24Fc-treated patients (pooled over time) showed a decline in cytokine correlations compared to baseline or placebo treatment. Similarly, cytokine network plots connecting cytokines with moderate and strong associations (Pearson correlation r>0·4^21^) showed lower overall interconnectedness in CD24Fc group as compared to baseline or placebo treatment (Fig 4G). The overall cytokine network correlations and connectivity in CD24Fc-treated patients were significantly different from baseline or placebo treatment (Fig 4H-I).

To understand the relevance of decreased correlation and connectivity of the cytokine network in CD24Fc treated patients to disease severity and therapeutic effect, we performed a similar analysis using a previously published dataset of cytokine expression in serum from patients with COVID-19 who were either treated in the intensive care unit (ICU patients) or did not require ICU treatment (non-ICU patients)^31^. Notably, we found that inter-cytokine correlation and connectivity was lower in non-ICU patients than ICU patients (**Fig S5**). These data suggest that increased blood cytokine network correlation and connectivity are associated with increased COVID-19 disease severity, while mild disease (without the need for ICU treatment) is characterized by lower correlation and connectivity. Therefore, decreased correlation and connectivity of the cytokine network in CD24Fc-treated patients are likely evidence of therapeutic efficacy.

To identify factors that may play an important role in response to CD24Fc, we calculated centrality scores^24^ for individual cytokines based on their connectivity and correlations within the global cytokine network (Table S4 and **S5**). The variances of the centrality scores of 30 cytokines were lower in baseline and placebo-treated COVID-19 patients compared to HD and CD24Fc-treated COVID-19 patients (Fig 4J). These data indicate that distinct cytokines are highly heterogeneous in terms of their interconnectedness with other cytokines (centrality) in healthy individuals. Upon SARS-CoV-2 infection, cytokine centralities become more uniform and subsequent CD24Fc treatment abrogates this effect (Fig 4J).

## DISCUSSION

Patients enrolled in the SAC-COVID clinical trial, a subpopulation of which were studied herein, demonstrated accelerated clinical recovery following CD24Fc treatment compared to placebo. CD24Fc was generally well-tolerated, reduced disease progression, and shortened hospital length of stay (results under review in Welker J *et al.* “Therapeutic Efficacy and Safety of CD24Fc in Hospitalised Patients with COVID-19,” submitted to *Lancet*). Given the proposed mechanism of action and pathophysiology of SARS-CoV-2, we hypothesized that CD24Fc reduced the hyperactive systemic immune responses in infected patients leading to accelerated return to homeostasis. Using deep immune profiling of longitudinal samples combined with our deep bioinformatic analysis, we uncovered the effects of CD24Fc on the systemic host immune response. Overall, we found that CD24Fc treatment blunted immune cell activation across several compartments and facilitated return to a more normal phenotype following SARS-CoV-2 infection.

Comparing baseline COVID-19 patients with HD allowed us to identify the immune cell populations driving pathogenesis. As expected, we saw a significant increase in activated CD8^+^ T and NK cells in SARS-CoV-2-infected patients. We augmented the unbiased clustering analysis with a semi-supervised approach to define an unbiased activation score. CD24Fc-treated patients demonstrated significant reduction in activation score over time for both CD8^+^ T and NK cells compared to placebo-treated patients.

The changes in population dynamics between HD and COVID-19 patients are intriguing and offer two separate interpretations. CD24Fc may preferentially block the differentiation of mature B cells into effector plasma cells, resulting in relatively fewer plasma B cells (cluster 6) and more mature B cells (cluster 8). Alternatively, CD24Fc treatment may reduce the systemic burden of SARS-CoV-2 infection, which would limit the number of plasma cells due to accelerated recovery. In either scenario, the correlation between decreased circulating plasma cells in CD24Fc-treated patient samples suggests significant immuno-modulatory roles in this treatment. The ability of patients to mount an effective anti-Spike antibody response was not compromised by CD24Fc treatment.

Aberrant and rapid increase in a broad spectrum of pro-inflammatory cytokines, known as a cytokine storm, plays a central role in pathogenesis of ARDS and other severe complications of SARS-CoV-2 infection^32^. Unlike the cytokine storm associated with immunotherapy, which can be effectively treated by antibodies targeting IL-6R, treating COVID-19 with the same antibodies has shown limited success. Our longitudinal analysis revealed a broad-spectrum up-regulation of systemic cytokines in patients with severe COVID-19. More importantly, CD24Fc treatment cause rapid and sustained reduction of most of the 30 cytokines/chemokines tested. Among them are known COVID-19 therapeutic targets such as IL-6 and GM-CSF. This broad effect may explain the significant therapeutic efficacy of CD24Fc in treating hospitalized COVID-19 patients.

In addition to the two therapeutic targets, we also identified two cytokines that were significantly downregulated after CD24Fc treatment: IL-10 and IL-15. Both are linked with COVID-19 severity, increased intensive care admission, and/or COVID-19-associated death^33–35^. Although generally associated with immunosuppressive functions, IL-10 can also stimulate NK and CD8^+^ T cells and induce B cell proliferation and antibody production^36^. IL-15 promotes activation and expansion of NK and CD8^+^ T cells^37, 38^. Thus, CD24Fc may blunt NK and CD8^+^ T cell activation by suppressing IL-10 and IL-15 production. Since IL-15 also promotes activation and recruitment of neutrophils to site of inflammation, CD24Fc may suppress COVID-19-associated neutrophil activation and/or neutrophilia^39^. Furthermore, CD24Fc may limit viral replication by suppressing IL-10 production, which has been shown to enhance viral replication of HIV, HCV and HBV^40^.

Importantly, unlike HD, COVID-19 patients displayed strong positive correlations between inflammatory cytokines, consistent with broad misfiring of host immune responses^29, 31, 41^. Notably, CD24Fc treatment reduced systemic cytokine levels and diminished correlations and connectivity in SARS-CoV-2-infected individuals, thus reshaping the systemic cytokine network towards a less tightly co-regulated state characteristic of homeostasis. Based on analysis of the global cytokine landscape, we conclude that CD24Fc mitigates the exacerbated host systemic inflammatory responses to SARS-CoV-2. This conclusion was corroborated by the decrease of cytokine correlation and connectivity in patients with mild COVID-19 infections as compared to patients with severe disease that required an ICU treatment. A detailed investigation of individual inflammatory markers revealed potential mechanisms of COVID-19 severity reduction by CD24Fc.

In conclusion, the data presented here offer unique immunological insights that underscore the clinical findings of the SAC-COVID trial. These results strongly support further investigation of CD24Fc for various inflammatory conditions including COVID-19. Our unique cytokine centrality analysis and cellular activation index also warrants further study as a prognostic tool for guiding therapy in COVID-19 and other systemic inflammatory conditions.

## Data Availability

The authors confirm that the data supporting the findings of this study are available within the article and its supplementary materials. Raw data that support the findings of this study are available on request from the corresponding author, Dr. Zihai Li

## Acknowledgements and Funding.

This work was supported in part by National Institutes of Health (NIH) grants, including R01AI077283, R01CA213290 to Z.L; and R37CA233770 to A.E.V. Z.L is also supposed by funding from the Pelotonia Foundation. Research reported in this publication was also supported by The Ohio State University Comprehensive Cancer Center and the NIH under grant number P30CA016058.

We acknowledge the patients who agreed to participate in this clinical trial, as the forward trajectory of science hinges on their support. We acknowledge Oncoimmune (now part of Merck & Co., Inc., Kenilworth, NJ, USA) for designing and supporting the clinical trial, which allowed us to collect samples for our correlative studies. The correlative studies were not part of the original phase III Oncoimmune trial, but were done at The Ohio State University trial site after communication of intent and sample collection. We thank the staff and researchers in the Pelotonia Institute for Immuno-Oncology for their support during the course of the study. We appreciate the expert administrate support by Ms. Teresa Kutcher.

## RESEARCH IN CONTEXT

### Evidence before this study

We searched Pubmed, medRxiv, and bioRxiv from January 2020 for publications that studied the efficacy of therapeutic reagents and their effect on immune cell population dynamics and T and NK cells activation status in COVID-19 patients. Search terms used broadly were: immune cell dynamics in COVID-19 patients, effects of COVID-19 therapeutic reagents on immune system, COVID-19 immuno-pathology, CD24Fc and COVID-19. To our knowledge, extensive immune phenotyping and investigation of T and NK cell activation status has not been done in a phase III clinical trial setting evaluating therapeutic efficacy of CD24Fc. Existing approaches for characterizing T and NK cell activation have failed to succinctly model activation status as a function of treatment group over the course of the disease, thus preventing robust conclusions regarding the effect of treatment on immune cell dynamics in COVID-19 patients.

### Added value of this study

In the SAC-COVID phase III clinical trial (NCT04317040), CD24Fc treatment accelerates recovery of patients with severe COVID-19 (clinical results under review in Welker J *et al*. “Therapeutic Efficacy and Safety of CD24Fc in Hospitalised Patients with COVID-19,” submitted to *Lancet*). CD24Fc reduced disease progression and shortened hospitalization, with no apparent side effects. In the current manuscript, we show that CD24Fc reduces abnormal systemic inflammation as a key mechanism of action. We developed a statistical framework for characterizing T and NK cell activation longitudinally using repeated flow cytometry samples taken at each study timepoint. Our method is generalized and may be applied to uncover disease dynamics in other studies, especially in the context of COVID-19. Even though vaccines significantly reduce morbidity and mortality associated with SARS-CoV-2 infection, dexamethasone is the only therapeutic modality currently known to effectively treat patients with severe COVID-19. The emergence of variants such as the Delta variant, which shows evidence of escaping immunoprotection conferred by vaccination, emphasizes the importance of continued investigation into drugs that will mitigate severe COVID-19 symptoms. Development of drugs targeting the excessive immune response characteristic of COVID-19 remains crucial, and the data presented here and by Welker at al. support the use of CD24Fc to reduce morbidity and mortality associated with severe COVID-19.

### Implications of all the available evidence

An excessive immune system activation upon SARS-CoV-2 infection can cause lethal complications and diseases progression mainly caused by cytokine storm. To counter this extreme immune responses, dexamethasone, well established anti-inflammatory reagent, has been used in clinic and showed its effectiveness in severe COVID-19 cases. CD24Fc, also showed therapeutic efficacy (Welker at al.) and here we studied potential mechanism of CD24Fc treatment. CD24Fc treatment systemically reduced activation of T and NK cells, and also reduced cytokines responsible for COVID-19 immunopathology. This suggests that CD24Fc can be used as a new therapeutic tool to treat COVID-19 patients.

## SUPPLEMENTAL MATERIAL

This appendix has been provided by the authors to give readers additional information about their work.

## LIST OF INVESTIGATORS

**Clinical Trial Site:** The Ohio State University Wexner Medical Center 410 W. Tenth Ave, Columbus OH 43210

**Clinical Laboratory Facility:** OSUCCC Pelotonia Institute for Immuno-Oncology, 460 W. 12th Ave Room 580 Biomedical Research Tower, Columbus OH 43210

**Clinical Investigators:** Carlos Diego Malvestutto, M.D. M.P.H; Zeinab El Boghdadly, M.D.; Mohammad Mahdee Sobhanie, M.D.; Jose Bazan, D.O.; Mark Lustberg, M.D. Ph.D.; Susan Koletar, M.D.; Zihai Li, M.D. Ph.D.; Kelsi Reynolds; Karthik Chakravathy

**Complete list of contributors (alphabetical):** Carter Allen^1,3,4^, Jose Bazan^2^, Chelsea Bolyard^3^, Zeinab El Boghdadly^2^, Donna Bucci^3^, Karthik B. Chakravarthy^3,11^, Yuzhou Chang^1,3,4^, Dongjun Chung^3,4^, Martin Devenport^13^, John P. Evans^12^, Manuja Gunasena^8,9^, Aastha Khatiwada^5^, Susan Koletar^2^, Amrendra Kumar^6,10^, Anqi Li^1,3,11^, Zihai Li^2,3^, Shan-Lu Liu^12^, Yang Liu^13^, Namal P. M. Liyanage^8,9^, Mark Lustberg^2^, Anjun Ma^4^, Qin Ma^4^, Carlos D. Malvestutto^2^, Kelsi Reynolds^3^, Brian P. Riesenberg^3^, Mohammad Mahdee Sobhanie^2^, No-Joon Song^3^, Zequn Sun^5^, Maria Velegraki^3^, Anna E. Vilgelm^3,6,10^, Kevin P. Weller^3^, Menglin Xu^2^, Mohamed Yusuf^6^, Cong Zeng^12^, Pan Zheng^13^.

**Affiliations:**

^1^The Ohio State University, Columbus, OH 43210, USA

^2^Department of Internal Medicine, The Ohio State University College of Medicine, Columbus, OH

^3^The Pelotonia Institute for Immuno-Oncology, The Ohio State University Comprehensive Cancer Center, Columbus, OH 43210, USA

^4^Department of Biomedical Informatics, The Ohio State University College of Medicine, Columbus, OH

^5^Department of Public Health Sciences, Medical University of South Carolina, Charleston, SC

^6^The Ohio State University Comprehensive Cancer Center, Columbus, OH 43210, USA

^7^Department of Microbiology, The Ohio State University College of Arts and Sciences, Columbus, OH 43210, USA ^8^Department of Microbial Infection and Immunity, The Ohio State University College of Medicine, Columbus, OH 43210, USA

^9^Department of Veterinary Biosciences, The Ohio State University College of Veterinary Medicine, Columbus, OH 43210, USA

^10^Department of Pathology, The Ohio State University College of Medicine, Columbus, OH

^11^The Ohio State University College of Medicine, Columbus, OH 43210, USA

^12^Center for Retrovirus Research and Department of Veterinary Biosciences, The Ohio State University, Columbus, OH 43210, USA

^13^OncoC4, Rockwille, MD, USA

**Author contributions are as follows (alphabetically):**

***Study conception and design:*** D Bucci, M Devenport, Z Li, SL Liu, Y Liu, C Malvestutto, NJ Song, P Zeng ***Acquisition of data:*** D Bucci, K Chakravarthy, JP Evans, M Gunasena, A Kumar, A Li, N Liyanage, C Malvestutto, K Reynolds, BP Riesenberg, NJ Song, M Velegraki, AE Vilgelm, K Weller, M Yusuf

***Analysis and interpretation of data:*** C Allen, Y Chang, D Chung, JP Evans, K Chakravarthy, A Khatiwada, A Kumar, A Li, Z Li, SL Liu, N Liyanage, A Ma, Q Ma, BP Riesenberg, NJ Song, Z Sun, M Velegraki, AE Vilgelm, K Weller, M Xu, C Zeng

***Drafting of manuscript:*** C Allen, C Bolyard, K Chakravarthy, D Chung, A Kumar, Z Li, SL Liu, N Liyanage, Q Ma, C Malvestutto, BP Riesenberg, NJ Song, AE Vilgelm, K Weller

Critical revision: C Allen, C Bolyard, D Chung, Z Li, SL Liu, BP Riesenberg, NJ Song

***Other:*** Z Li, overall supervision of study activities; AE Vilgelm, acquisition of funding and supervision of cytokine studies; M Devenport, Y Liu, P Zheng for clinical study.

## SUPPLEMENTARY METHODS

### PATIENTS AND TRIAL PROCEDURE

This study included samples from patients enrolled in NCT04317040 at The Ohio State University Wexner Medical Center. Patients eligible for this trial were hospitalized with COVID-19, requiring supplemental oxygen but not mechanical ventilation, and had a prior positive SARS-CoV-2 PCR test. Enrolled patients were randomized in a double-blinded fashion by the hospital pharmacist to receive either a single dose of CD24Fc antibody (480mg IV infusion) or placebo control (IV saline). Peripheral blood samples were collected from patients prior to drug infusion (D1), and at subsequent time points 1, 3, 7, 14, and 28 days after drug infusion (D2, D4, D8, D15, and D29). Patients were monitored until D29, after which they completed the study endpoint. Pertinent patient clinical information was abstracted from the internal electronic medical record database including demographic data, medical history, clinical laboratory findings, and treatment regimen for COVID-19 during hospital stay (Table S1). All enrolled patients were able to complete the study endpoint with no demises in either group. After enrollment and completion of the study period, two patients were excluded from analysis. One exclusion was due to a diagnosis of chronic lymphocytic leukemia (CLL) which confounded the subsequent immunological analyses. Another exclusion occurred with a patient who received an infusion but was discharged before any post-infusion peripheral blood sample could be collected; hence no comparative analysis could be made using this patient. Written or witnessed oral informed consent was obtained for each patient. This trial and protocol were approved by Western Institutional Review Board. The study was monitored continuously by a clinical monitor and a medical monitor from the contract research organization (CRO) who also generated safety reports submitted to an independent Data and Safety Monitoring Board (DSMB). Data quality control checks were performed and medical monitor verified that the clinical trial was conducted and data was generated in compliance with protocol, International Conference on Harmonization Good Clinical Practice (ICH GCP) and all applicable regulatory requirements.

Patient characteristics were clinically matched between the two groups. All patients enrolled in the study received a treatment regimen for COVID-19 by hospital care teams regardless of their placebo/CD24Fc treatment status. Patients were randomized in a double-blind fashion into CD24Fc antibody treatment group (n=10) or placebo control group (n=12).

### PBMC COLLECTION AND FLOW CYTOMETRY STAINING

Samples for this study were collected from patients enrolled in clinical trial NCT04317040. We analyzed samples from 22 patients hospitalized at The Ohio State University Wexner Medical Center with severe COVID-19. Peripheral blood mononuclear cells (PBMCs) were isolated per manufacturer’s protocol using CPT tubes (BD Bioscience). Healthy donor (HD) PMBCs were obtained from STEMCELL Technologies™. We utilized a 36-color flow cytometry panel (Table S2, developed by Cytek^1^) to distinguish immune populations; we developed a 25-color panel (Table S2) to study activation status of CD8^+^, CD4^+^, and CD56^+^ subsets. For the 25-color panel, surface markers were stained in 4°C for 1h and FOXP3/Transcription Factor Staining Buffer Set (eBioscience™) was used per manufacturers recommendation to perform intracellular staining. Cells were analyzed using the Cytek Aurora system.

### VIRUS NEUTRALIZATION ASSAY

Virus was produced as previously described^2^, and incubated with COVID- 19 patient sera for 1h at 37°C. Virus was then overlaid onto ACE2-expressing 293T cells for 6h. Gluc activity was measured 24, 48, and 72 hours after infection.

### CYTOKINE AND CHEMOKINE ASSAY

Plasma samples were processed using multiplexed ELISA-based platform Quantibody® Human Inflammation Array 3 (RayBiotech QAH-INF-3) in accordance with manufacturer’s protocol. Slides were shipped to manufacturer site for scanning and data extraction services. Raw optical data were analyzed using manufacturer’s analysis tool to construct standard curves and determine absolute cytokine concentrations. Cytokines for which standards did not yield good standard curve fit or that were undetectable were excluded (IFNγ, IL1rα, IL2, IL13, MCP-1, TNFα, TNFβ, IL-11, IL-12p70, IL-17A). Seven of these cytokines were detected using an alternative method. Specifically, cytokines IFNγ, IL1rα, IL2, IL13, MCP-1, TNFα, and IL-12p70 were measured by Luminex analysis. For that, plasma samples were sent to EVE Technologies that performed the assay and provided cytokine concentration data (Table S6).

### FLOW CYTOMETRY DATA ANALYSIS

We integrated flow cytometry marker data from all samples and arcsinh scaling was applied using OMIQ (https://www.omiq.ai/). Then, we visualized cells in a reduced two-dimensional space using the UMAP algorithm implemented in the R package uwot^3^. We adopted a multivariate t-mixture model to cluster cells based on the normalized multivariate flow cytometry marker expression^4^. For each data set, we chose the optimal number of cell clusters by selecting the model with the minimum Bayesian information criterion (BIC) score^5^. Then, we annotated cell types by visually investigating heatmaps of median marker expressions across clusters and expressions of these markers on the UMAP space.

### IMMUNE CELL ACTIVATION SCORE CONSTRUCTION

To measure activation, we defined a cell-level immune cell activation score for each flow cytometry data set. We selected a subset of immune cell activation markers from the panel^6, 7^, and ran a principal component analysis (PCA) comparing cells from HD and baseline (Day 1) COVID patients, using these activation markers as features. We used the first principal component (PC1) as an activation score to reflect the differences in immune cell activation between groups. The loadings of each pre-selected activation marker onto PC1 were used as coefficients to compute an activation score for COVID-19 patients after baseline.

### CYTOKINE SCORE CONSTRUCTION

To construct the cytokine score, we implemented a weighted sum approach, motivated by the polygenic risk score calculation in the genome-wide association study (GWAS). First, we fit a generalized linear mixed model (GLMM) of each cytokine measurement (base 10 log-transformed) on treatment, time, treatment*time, age, sex, and race as fixed-effect terms, along with subject-level random effect terms. Second, the *p*-value for evaluating the overall difference in trends between CD24Fc and placebo groups across all the time points was calculated using the Kenward-Roger method^8^. Finally, we obtained the weighted sum of cytokine measurements using the -2 log transformed *p*-value for the trend difference as weights, motivated by the Fisher’s method. We validated the above approach using the PCA and autoencoder approaches^9^.

### NETWORK-LEVEL ANALYSIS OF CYTOKINE DATA

We first calculated Pearson correlation coefficients between cytokines (base 10 log-transformed). Then, we constructed a network, where a node represents a cytokine and an edge between two nodes was built if the corresponding absolute correlation coefficient is larger than 0.4, a cutoff that is usually considered to be moderate correlation^10^. The weight of an edge represents the corresponding correlation coefficient. A network was built via the MetScape^11^ (version 3·1·3) application in Cytoscape^12^ (version 3·8·0). We evaluated the network structure and the importance of each node in the network based on an eigenvector centrality (EC) score^13^ using the CytoNCA^14^ (version 2·1·6) application in Cytoscape (version 3.8.0). Nodes with larger EC scores can be considered of higher importance.

### STATISTICAL ANALYSIS

All data were analyzed using the R statistical package. Group comparisons were evaluated using independent sample t-test or Kruskal-Wallis test for continuous variables, and Chi-squared test for categorical variables. In the longitudinal analyses, the overall differences in trends between CD24Fc and placebo groups across all the time points were evaluated using a GLMM of each measurement on treatment, time, treatment*time, age, sex, and race as fixed-effect terms, along with patient-level random intercepts. All mixed models were fit using the lme4 package^15^. The p-value for evaluating the overall difference in trends between CD24Fc and placebo groups across all the time points was calculated using the Kenward-Roger method^8^. The observed values and trend lines are centered at the baseline.

### TREATMENT GROUP DETERMINATION

The treatment group (control vs. CD24Fc) was determined by the post-infusion sera to absorb anti-CD24 antibody for staining of human CD24^+^ cells by flow cytometry. Patient group on the CD24Fc arm was further confirmed using CD24Fc ELISA (capture antibody: purified anti-human CD24, Clone ML5, BD bioscience, Cat#555426. San Jose, CA).

## SUPPLEMENTARY FIGURES

**Figure S1.**
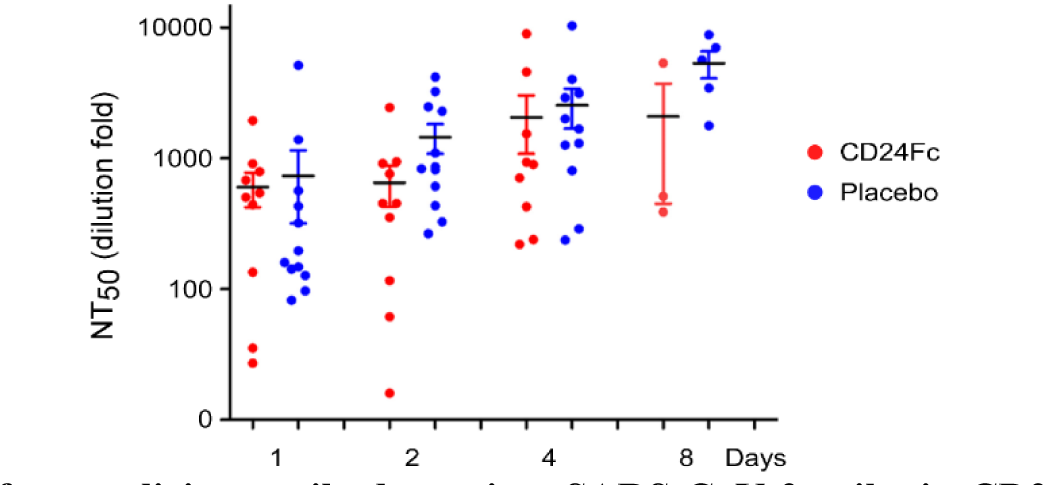
Comparison of neutralizing antibody against SARS-CoV-2 spike in CD24Fc treated patients and placebo group. Using our secreted nano luciferase-bearing pseudotyped lentivirus virus neutralization assay, we assessed the neutralizing antibody (nAb) titers for the CD24Fc treated and placebo groups throughout their treatment period. The average 50% neutralization titer (NT50) for both groups show an increase in antibody titers from day 0 to day 15, but no significant differences were observed when CD24Fc group were compared to placebo group.

**Figure S2.**
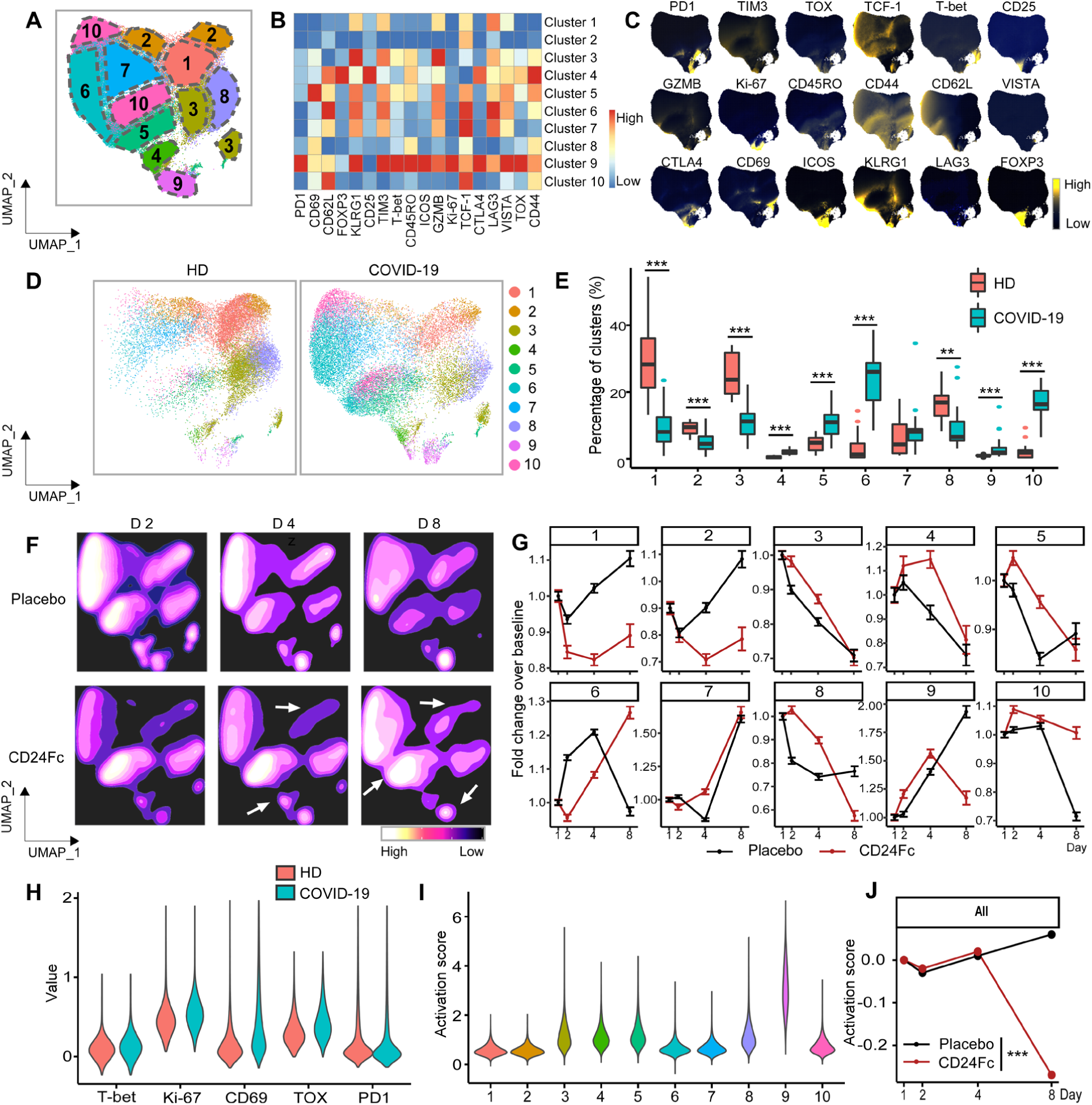
Subcluster analysis of peripheral blood CD4^+^ T cells in COVID-19 patients: activation following SARS-CoV2 infection is dampened by CD24Fc treatment. We clustered 1,203,034 CD4^+^ cells from HD (n=17) and COVID-19 (n=22) patients using an unbiased multivariate *t*- mixture model, which identified 10 CD4^+^ sub-clusters that reflect statistically distinct cell activation states. We visualized the relative similarity of each cell and cell cluster on the two-dimensional UMAP space with a 10% downsampling (**Panel A**). Using median expression of flow cytometry markers, we generated a cluster-by-marker heatmap to characterize the subsets (**Panel B**) and visualized individual marker expression patterns on the UMAP space (**Panel C**). To understand the effect of SARS-CoV2 infection on cell population dynamics, we compared UMAP dot plots (**Panel D**) and cluster frequencies (**Panel E**) of HD vs. baseline COVID-19 patient samples (cluster 1, p<0·001; cluster 2, p<0·001; cluster 3, p<0·001; cluster 4, p<0·001; cluster 5, p<0·001; cluster 6, p<0·001; cluster 8, p=0·002; cluster 9, p<0·001; cluster 10, p<0·001). We visualized samples from COVID-19 patients D2, 4, and 8 after CD24Fc vs. placebo treatment using contour plots to represent the density of cells throughout regions of the UMAP space (**Panel F**). We describe cluster population dynamics as fold change over baseline in each treatment group (**Panel G**; sample distribution described in Fig 1F legend). To better characterize the activation status of CD4 T cells, we linearly transformed a subset of markers (T-bet, Ki-67, CD69, TOX, PD1) to create a univariate cell-level activation score (**Panel H**), where highly activated cell clusters (such as cluster 9) had highest activation scores (**Panel I**). We then fit a GLMM to our longitudinal cell-level activation scores to assess the effect of CD24Fc treatment on activation scores over time (**Panel J**; p<0·001). The p-value for evaluating the overall difference in trends between CD24Fc and placebo groups across all time points was calculated using the Kenward-Roger method. Using this model, we found that CD24Fc-treated samples had significantly lower CD4^+^ cell activation levels relative to placebo. **\*\***, p<0·01; **\*\*\***, p<0·001.

**Figure S3.**
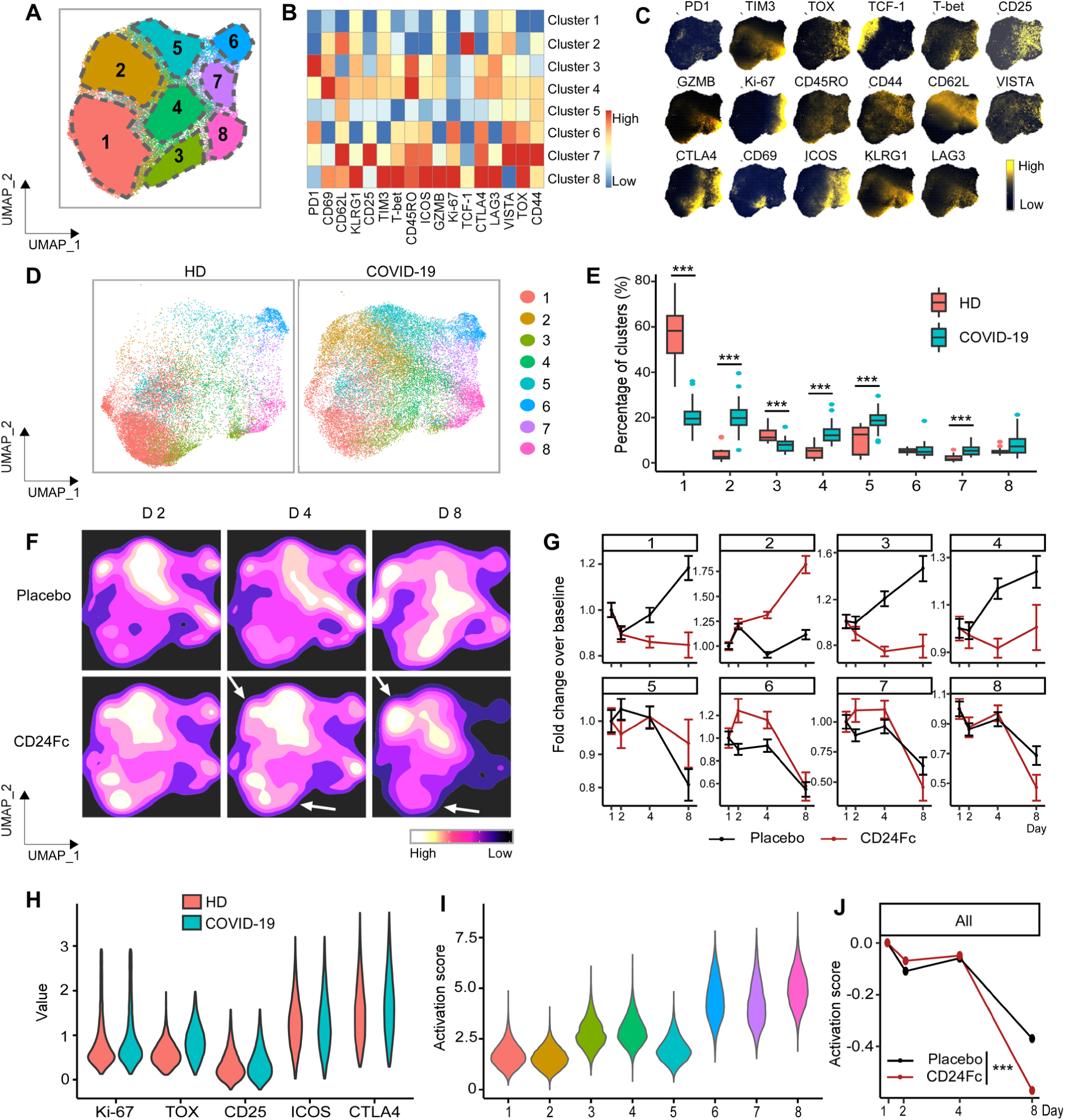
Subcluster analysis of peripheral blood FOXP3^+^ Treg cells in COVID-19 patients: activation following SARS-CoV2 infection is dampened by CD24Fc treatment. We clustered 98,525 FOXP3+ Treg cells from HD (n=17) and COVID-19 (n=22) patients using an unbiased multivariate *t*-mixture model, which identified 8 FOXP3^+^ Treg sub-clusters that reflect statistically distinct cell activation states. We visualized the relative similarity of each cell and cell cluster on the two-dimensional UMAP space with a 10% downsampling (**Panel A**). Using median expression of flow cytometry markers, we generated a cluster-by-marker heatmap to characterize the subsets (**Panel B**) and visualized individual marker expression patterns on the UMAP space (**Panel C**). To understand the effect of SARS-CoV2 infection on cell population dynamics, we compared UMAP cluster frequencies of HD vs. baseline COVID-19 patient samples (**Panels D** and **E**). We visualized samples from COVID-19 patients D2, 4, and 8 after CD24Fc vs. placebo treatment using contour plots to represent the density of cells throughout regions of the UMAP space (**Panel F**). We describe cluster population dynamics as fold change over baseline in each treatment group (**Panel G**; sample distribution described in Fig 1F legend). To better characterize the activation status of Treg cells, we linearly transformed a subset of markers (Ki-67, TOX, CD25, ICOS, CTLA4) to create a univariate cell-level activation score (**Panel H**), where highly activated cell clusters (such as clusters 6, 7 and 8) had highest activation scores (**Panel I**). We then fit a GLMM to our longitudinal cell-level activation scores to assess the effect of CD24Fc treatment on activation scores over time (**Panel J**). The p-value for evaluating the overall difference in trends between CD24Fc and placebo groups across all time points was calculated using the Kenward-Roger method. Using this model, we found that CD24Fc-treated samples had significantly lower Treg cell activation levels relative to placebo.

**Figure S4.**
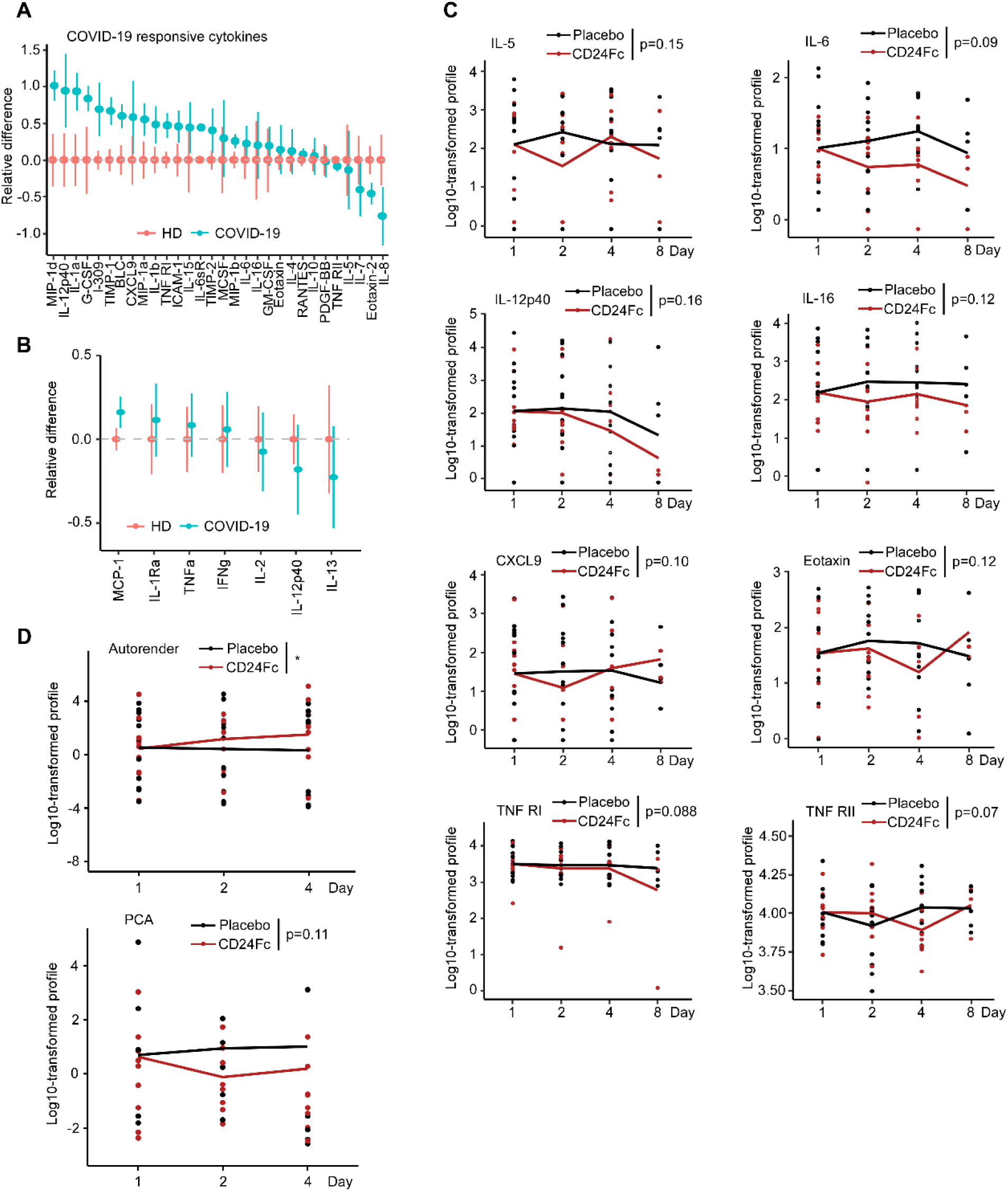
CD24Fc treatment downregulates systemic cytokines response in patients with COVID-19. We studied plasma cytokine and chemokine levels in HD and COVID-19 patients. Cytokine/chemokine measurements were log-transformed, and relative differences in cytokines in COVID-19 (n=22) compared to HD (n=25) samples were depicted (**Panel A and B**). Graph in Panel A shows data obtained using multiplex-ELISA platform, while graph in panel B presents data of cytokines measured by the Luminex analysis. Independent sample t-test was used to evaluate equality of average cytokine/chemokine levels. A number of other markers displayed trends towards decline in CD24Fc cohort compare to placebo, although these changes were not statistically significant (**Panel C**). Log-10 transformed cytokine measurement (dots) and GLMM predicted fixed effects trends (lines) of IL-5, IL-6, IL-12p40, IL-16, CXCL9, Eotaxin, TNF R1 and TNF RII plasma concentrations in CD24Fc (red) and placebo (black) groups are displayed. The observed values and trend lines are centered at D1 mean. Longitudinal analysis of cytokine score was confirmed using both Autoencoder and PCA approaches (**Panel D**). We applied PCA and autoencoder on the base 10 log-transformed, centered and scaled cytokine data, and investigated the first two principal components (PCs) from the PCA and the three latent components from the autoencoder as cytokine scores. The autoencoder analysis was implemented using the Keras package. Specifically, we set one hidden layer for encoder and decoder, respectively, and three-dimensional embedding as latent layer output. All parameters were trained based on a 3-fold cross-validation. Due to missing data on D8, only D1, D2, and D4 data were used for the cytokine score calculation. For **Panels C** and **D**, the overall differences in trends between CD24Fc and placebo groups across all the time points were evaluated using a GLMM of each measurement. The p-value for evaluating the overall difference in trends between CD24Fc and placebo groups across all the time points was calculated using the Kenward-Roger method.

**Figure S5.**
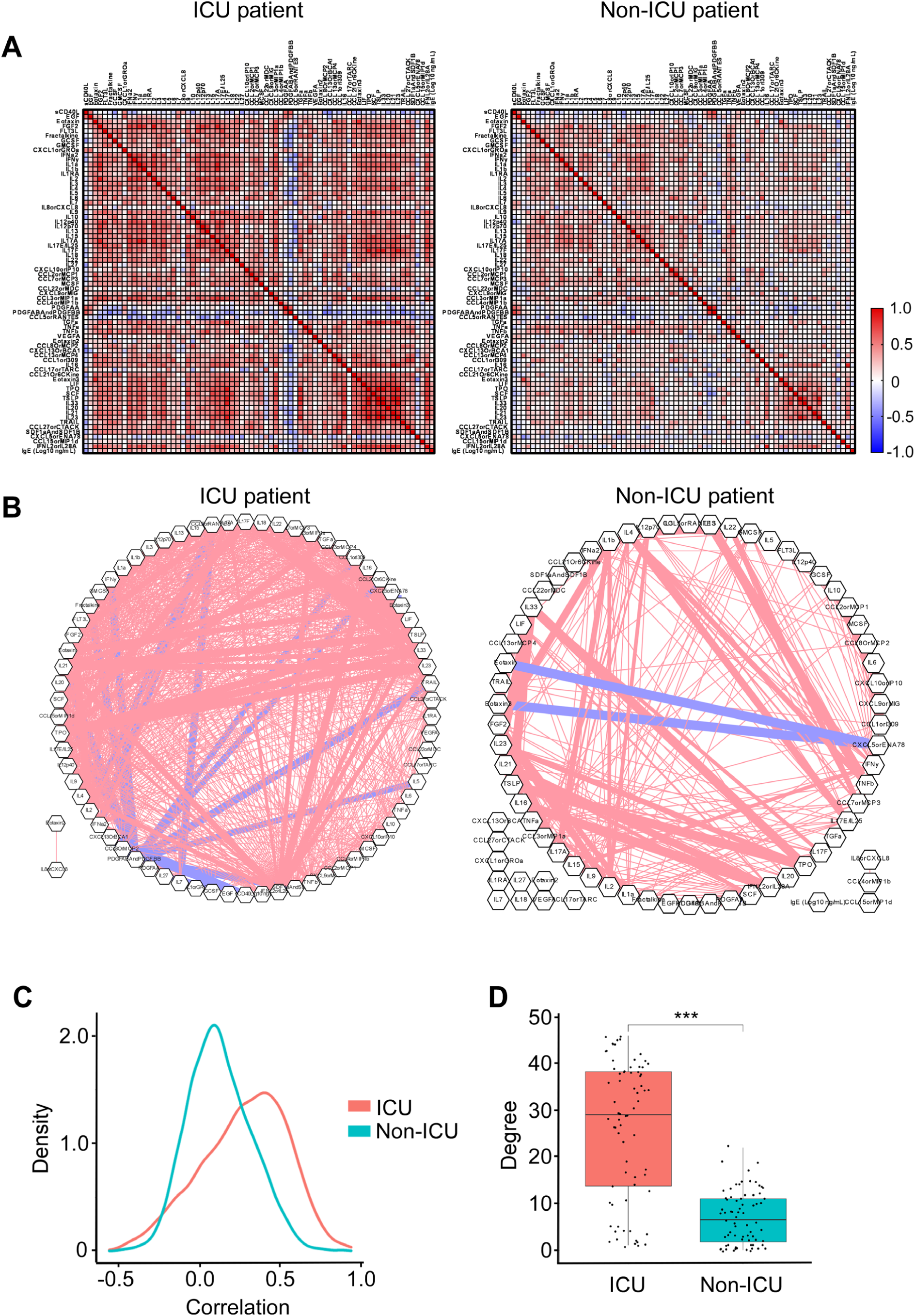
Patients with severe COVID-19 that require an ICU treatment display increased correlation and connectivity of the systemic cytokine network. We analyzed correlation (**Panel A**) and connectivity (**Panel B**) between circulating cytokines and chemokines in COVID-19 patients that either required (ICU patients), or did not require an ICU treatment (non-ICU patients). Cytokine measurements were obtained from previously published dataset ^16^. Analysis was performed as described in Fig 4. A density plot constructed based on connectivity between plasma cytokines is shown in **Panel C**. **Panel D** shows an association between the severity of COVID-19 infection and the degree of the connectivity between plasma cytokines with severe UCU cases displaying higher degree of connectivity. The p-value was calculated using Wilcoxon Rank Sum test.

## SUPPLEMENTARY TABLES

**Table S1.** Patient Characteristics.

| CHARACTERISTIC | OVERALL<br>(N=22) | PLACEBO<br>(N=12) | CD24Fc<br>(N=10) | P-value |
| --- | --- | --- | --- | --- |
| **a) Demographics** |  |  |  |  |
| BMI - median (IQR) | 31.65 (28.18-38.83) | 31.65 (29.41-39.52) | 32.2 (28.1-37.88) | 0.644 |
| Age, yr - median (IQR) | 57 (50.25-74.75) | 60.5 (53.5-75) | 55 (49.5-62) | 0.62 |
| <65 yr - no. (%) | 15 (68.2) | 7 (58.3) | 8 (80.0) |  |
| ≥65 yr - no. (%) | 7 (31.8) | 5 (41.7) | 2 (20.0) |  |
| Sex - no. (%) |  |  |  | 0.903 |
| Female | 8 (36.4) | 5 (41.7) | 3 (30.0) |  |
| Male | 14 (63.6) | 7 (58.3) | 7 (70.0) |  |
| Race - no. (%) |  |  |  | 0.528 |
| White | 16 (72.7) | 9 (75.0) | 7 (70.0) |  |
| Black/AA | 5 (22.7) | 2 (16.7) | 3 (30.0) |  |
| Not Specified | 1 (4.5) | 1 (8.3) | 0 (0.0) |  |
| Ethnicity - no. (%) |  |  |  | 0.724 |
| Hispanic | 4 (18.2) | 3 (25.0) | 1 (10.0) |  |
| Non-Hispanic | 18 (81.8) | 9 (75.0) | 9 (90.0) |  |
| Smoking Hx - no. (%) | 7 (31.8) | 3 (25.0) | 4 (40.0) | 0.77 |
| **b) Co-existing Conditions** |  |  |  |  |
| Comorbidities - no. (%) |  |  |  |  |
| Obesity (BMI ≥30) | 12 (54.5) | 7 (58.3) | 5 (50.0) | 1 |
| Hypertension | 11 (50.0) | 5 (41.7) | 6 (60.0) | 0.669 |
| Hyperlipidemia | 8 (36.4) | 5 (41.7) | 3 (30.0) | 0.903 |
| Heart Disease | 7 (31.8) | 2 (16.7) | 5 (50.0) | 0.226 |
| Diabetes | 7 (31.8) | 3 (25.0) | 4 (40.0) | 0.77 |
| Autoimmune Condition | 3 (13.6) | 3 (25.0) | 0 (0.0) | 0.281 |
| Cancer | 2 (9.1) | 1 (8.3) | 1 (10.0) | 1 |
| HIV | 1 (4.5) | 0 (0.0) | 1 (10.0) | 0.926 |
| COPD/Asthma | 2 (9.1) | 1 (8.3) | 1 (10.0) | 1 |
| **c) Clinical Information** |  |  |  |  |
| Baseline diastolic blood pressure, mm Hg - median (IQR) | 65.5 (61.5-74) | 67.5 (62.5-72) | 65.5 (61.75-73.25) | 0.766 |
| Baseline systolic blood pressure, mm Hg - median (IQR) | 127 (121-139.75) | 138.5 (117.5-144) | 124 (121.75-129) | 0.137 |
| Normotensive (<130) - no. (%) | 12 (54.5) | 5 (41.7) | 7 (70.0) | 0.369 |
| Hypertensive (≥130) - no. (%) | 10 (45.5) | 7 (58.3) | 3 (30.0) |  |
| Baseline O<sub>2</sub> saturation on room air, % - median (IQR) | 0.88 (0.84-0.9) | 0.86 (0.8-0.88) | 0.88 (0.85-0.9) | 0.185 |
| Hypoxic (<90) - no. (%) | 16 (72.7) | 10 (83.3) | 6 (60.0) | 0.458 |
| Non-hypoxic (≥90) - no. (%) | 6 (27.3) | 2 (16.7) | 4 (40.0) |  |
| Baseline respiratory rate, respirations/min - median (IQR) | 20 (18-25.5) | 22 (19.5-28.5) | 19 (16.5-21.5) | 0.053 |
| Eupnic (≤20) - no. (%) | 13 (59.1) | 6 (50.0) | 7 (70.0) | 0.607 |
| Tachypnic (>20) - no. (%) | 9 (40.9) | 6 (50.0) | 3 (30.0) |  |
| Baseline heart rate, beats/min - median (IQR) | 82.5 (67-91.75) | 82.5 (73-91.25) | 77.5 (66.25-92.5) | 0.817 |
| Eucardic (≤100) - no. (%) | 22 (100.0) | 12 (100.0) | 10 (100.0) | n/a |
| Tachycardic (>100) - no. (%) | 0 (0.0) | 0 (0.0) | 0 (0.0) |  |
| Baseline temperature, °C - median (IQR) | 37 (36.8-37.38) | 36.95 (36.77-37.12) | 37.15 (36.82-37.55) | 0.466 |
| Febrile (>37.0) - no. (%) | 9 (40.9) | 4 (33.3) | 5 (50.0) | 0.722 |
| Non-febrile (≤37.0) - no. (%) | 13 (59.1) | 8 (66.7) | 5 (50.0) |  |
| Baseline RBC Count, M/μL - median (IQR) | 4.61 (4.36-4.94) | 4.61 (4.26-4.94) | 4.62 (4.44-4.94) | 0.598 |
| Low (<4.3) - no. (%) | 4 (18.2) | 3 (25.0) | 1 (10.0) | 0.541 |
| Normal (4.3-5.5) - no. (%) | 15 (68.2) | 8 (66.7) | 7 (70.0) |  |
| Elevated (>5.5) - no. (%) | 3 (13.6) | 1 (8.3) | 2 (20.0) |  |
| Baseline WBC Count, K/μL - median (IQR) | 5.75 (5.23-8.07) | 6.79 (5.34-7.98) | 5.62 (4.85-8.78) | 0.429 |
| Low (<4.5) - no. (%) | 2 (9.1) | 0 (0.0) | 2 (20.0) | 0.124 |
| Normal (4.5-11.0) - no. (%) | 19 (86.4) | 12 (100.0) | 7 (70.0) |  |
| Elevated (>11.0) - no. (%) | 1 (4.5) | 0 (0.0) | 1 (10.0) |  |
| Baseline Neutrophils, K/μL - median (IQR) | 4.43 (3.9-6.8) | 4.93 (4.3-6.38) | 4.03 (3.15-6.75) | 0.323 |
| Baseline Lymphocytes, K/μL - median (IQR) | 1.04 (0.76-1.33) | 1.14 (0.69-1.6) | 0.92 (0.85-1.21) | 0.921 |
| Baseline Monocytes, K/μL - median (IQR) | 0.36 (0.29-0.57) | 0.32 (0.25-0.5) | 0.38 (0.34-0.57) | 0.235 |
| Baseline Eosinophils, K/μL - median (IQR) | 0.04 (0.04-0.06) | 0.04 (0.04-0.04) | 0.04 (0.04-0.06) | 0.129 |
| Baseline Basophils, K/μL - median (IQR) | 0.04 (0.04-0.04) | 0.04 (0.04-0.04) | 0.04 (0.04-0.04) | 0.097 |
| Baseline Hemoglobin, g/dL - median (IQR) | 13.4 (12.95-14) | 13.3 (12.7-14.15) | 13.45 (13.17-13.67) | 0.575 |
| Low (<13.2) - no. (%) | 8 (36.4) | 5 (41.7) | 3 (30.0) | 0.263 |
| Normal (13.2-16.4) - no. (%) | 12 (54.5) | 5 (41.7) | 7 (70.0) |  |
| Elevated (>16.4) - no. (%) | 2 (9.1) | 2 (16.7) | 0 (0.0) |  |
| Baseline Platelet Count, K/μL - median (IQR) | 224.5 (187.75-253) | 224 (171.75-251) | 224.5 (208.25-268.75) | 0.621 |
| Normal (150-450) - no. (%) | 21 (95.5) | 11 (91.7) | 10 (100.0) | 1 |
| Elevated (>450) - no. (%) | 1 (4.5) | 1 (8.3) | 0 (0.0) |  |
| Baseline D-dimer, μg/mL - median (IQR) | 0.95 (0.58-1.92) | 1.12 (0.69-1.65) | 0.79 (0.45-1.82) | 0.276 |
| Normal (<0.50) - no. (%) | 5 (22.7) | 1 (8.3) | 4 (40.0) | 0.21 |
| Elevated (≥0.50) - no. (%) | 17 (77.3) | 11 (91.7) | 6 (60.0) | |
| Baseline International Normalized Ratio, sec - median (IQR) | 1.1 (1-1.1) | 1.1 (1-1.15) | 1.1 (1-1.1) | 0.487 |
| Normal (0.9-1.1) - no. (%) | 17 (77.3) | 9 (75.0) | 8 (80.0) | 1 |
| Elevated (>1.1) - no. (%) | 5 (22.7) | 3 (25.0) | 2 (20.0) |  |
| Baseline ESR, mm/hr - median (IQR) | 51.5 (40-71) | 43 (35.75-61.5) | 64.5 (43.75-71) | 0.198 |
| Baseline CRP, mg/L - median (IQR) | 80.59 (68.91-142.69) | 80.59 (67.83-147.37) | 90.22 (72.53-132.12) | 0.895 |
| Baseline Troponin, ng/mL - median (IQR) | 0.01 (0.01-0.02) | 0.01 (0.01-0.01) | 0.01 (0.01-0.03) | 0.193 |
| Time from symptom onset to infusion, days - median (IQR) | 10.5 (8.25-12.75) | 10.5 (8.75-13) | 10.5 (8.25-11) | 0.571 |
| Earlier (≤10) - no. (%) | 11 (50.0) | 6 (50.0) | 5 (50.0) | 1 |
| Later (>10) - no. (%) | 11 (50.0) | 6 (50.0) | 5 (50.0) |  |
| Time from infusion to discharge, days - median (IQR) | 6 (4-8.75) | 6 (3.75-9) | 5.5 (4-7.5) | 0.618 |
| Shorter (≤7) - no. (%) | 15 (68.2) | 8 (66.7) | 7 (70.0) | 1 |
| Longer (>7) - no. (%) | 7 (31.8) | 4 (33.3) | 3 (30.0) |  |
| Total hospital stay, days - median (IQR) | 9 (6-11.75) | 9.5 (7.5-12.25) | 7 (6-10) | 0.371 |
| Shorter (≤10) - no. (%) | 15 (68.2) | 7 (58.3) | 8 (80.0) | 0.531 |
| Longer (>10) - no. (%) | 7 (31.8) | 5 (41.7) | 2 (20.0) |  |
| O<sub>2</sub> requirement at admission, L/min - median (IQR) | 2 (2-4) | 2 (2-8) | 2.5 (2-3.25) | 0.601 |
| None (<1) - no. (%) | 5 (22.7) | 3 (25.0) | 2 (20.0) | 0.598 |
| Low (1-49) - no. (%) | 16 (72.7) | 8 (66.7) | 8 (80.0) |  |
| High (≥50) - no. (%) | 1 (4.5) | 1 (8.3) | 0 (0.0) | |
| Peak O<sub>2</sub> requirement during hospital stay, L/min - median (IQR) | 6.5 (3.25-11.25) | 8.5 (4.5-26.25) | 5 (3.25-6.75) | 0.119 |
| Low (1-49) - no. (%) | 18 (81.8) | 9 (75.0) | 9 (90.0) | 0.724 |
| High (≥50) - no. (%) | 4 (18.2) | 3 (25.0) | 1 (10.0) | |
| O<sub>2</sub> requirement at discharge, L/min - median (IQR) | 3 (3-3) | 3 (3-3) | 3 (2-3) | 0.414 |
| None (<1) - no. (%) | 17 (77.3) | 10 (83.3) | 7 (70.0) | 0.816 |
| Low (1-49) - no. (%) | 5 (22.7) | 2 (16.7) | 3 (30.0) |  |
| ICU Stay - no. (%) | 5 (22.7) | 4 (33.3) | 1 (10.0) | 0.43 |
| **d) Concomitant Medication** |  |  |  |  |
| Concurrent COVID-19 Treatments - no. (%) |  |  |  |  |
| Convalescent Plasma | 19 (86.4) | 10 (83.3) | 9 (90.0) | 1 |
| Remdesivir | 19 (86.4) | 11 (91.7) | 8 (80.0) | 0.865 |
| Dexamethasone | 15 (68.2) | 9 (75.0) | 6 (60.0) | 0.77 |
| Anti-microbials | 16 (72.7) | 9 (75.0) | 7 (70.0) | 1 |

Median and Inter-quartile range (IQR) was determined for all continuous variables. P-values were obtained using Kruskal Wallis test continuous variables. Chi-square test was used to obtain p-values for categorical variables.

**Table S2.** Immune Cell Marker Panels.

| **Cytek Flow Cytometry Panel** |  |
| --- | --- |
| **Marker** | **Description** |
| CD45RA | BUV395 Mouse Anti-Human CD45RA |
| Viability dye | LIVE/DEAD™ Fixable Blue Dead Cell Stain Kit, for UV excitation |
| CD16 | BUV496 Mouse Anti-Human CD16 |
| CCR5 | BUV563 Mouse Anti-Human CD195 (CCR5) |
| CD11c | BUV661 Mouse Anti-Human CD11c |
| CD56 | BUV737 Mouse Anti-Human CD56 |
| CD8 | BD Horizon™ BUV805 Mouse Anti-Human CD8 |
| CCR7 | Brilliant Violet 421™ anti-human CD197 (CCR7) Antibody |
| CD123 | CD123 Monoclonal Antibody (6H6), Super Bright 436, eBioscience™ |
| CD161 | CD161 Monoclonal Antibody (HP-3G10), eFluor 450, eBioscience™ |
| IgD | BV480 Mouse Anti-Human IgD |
| CD3 | Brilliant Violet 510™ anti-human CD3 Antibody |
| CD20 | CD20 Monoclonal Antibody (HI47), Pacific Orange |
| IgM | Brilliant Violet 570™ anti-human IgM Antibody |
| IgG | BD Horizon™ BV605 Mouse Anti-Human IgG |
| CD28 | Brilliant Violet 650™ anti-human CD28 Antibody (clone CD28.2) |
| CCR6 | Brilliant Violet 711™ anti-human CD196 (CCR6) Antibody |
| CXCR5 | BV750 Rat Anti-Human CXCR5 (CD185) |
| PD-1 | Brilliant Violet 785™ anti-human CD279 (PD-1) Antibody |
| CD141 | BD Horizon™ BB515 Mouse Anti-Human CD141 |
| CD57 | FITC anti-human CD57 Antibody |
| CD14 | Spark Blue™ 550 anti-human CD14 Antibody |
| CD45 | CD45 Monoclonal Antibody (H130), PerCP |
| CD11b | PerCP/Cyanine5.5 anti-human CD11b Antibody |
| TCR_gd | TCR gamma/delta Monoclonal Antibody (B1.1), PerCP-eFluor 710, eBioscience™ |
| CD25 | CD25 Monoclonal Antibody (BC96), PE, eBioscience™ |
| CD4 | eFluor 568 Anti-human CD4 |
| CD24 | CD24 Monoclonal Antibody (eBioSN3 (SN3 A5-2H10)), PE-eFluor 610, eBioscience™ |
| CD95 | CD95 (APO-1/Fas) Monoclonal Antibody (DX2), PE-Cyanine5, eBioscience™ |
| CXCR3 | CD183 (CXCR3) Monoclonal Antibody (CEW33D), PE-Cyanine7, eBioscience™ |
| CD27 | CD27 Monoclonal Antibody (O323), APC, eBioscience™ |
| CD1c | Alexa Fluor® 647 anti-human CD1c Antibody |
| CD19 | Spark NIR™ 685 anti-human CD19 Antibody |
| CD127 | APC-R700 Mouse Anti-Human CD127 |
| HLA-DR | HLA-DR Monoclonal Antibody (L243), APC-eFluor 780, eBioscience™ |
| CD38 | CD38 APC-Fire810 |
| **Immune Monitoring Cytometry Panel** |  |
| **Marker** | **Description** |
| Viability dye | LIVE/DEAD™ Fixable Blue Dead Cell Stain Kit, for UV excitation |
| CD45 | CD45 Monoclonal Antibody (2D1), Super Bright 645, eBioscience™ |
| CD3 | BUV395 Mouse Anti-Human CD3 Clone SK7 |
| CD8 | CD8a Monoclonal Antibody (OKT8 (OKT-8)), Super Bright 436, eBioscience™ |
| CD4 | CD4 Monoclonal Antibody (RPA-T4), PerCP-Cyanine5.5, eBioscience™ |
| FOXP3 | FOXP3 Monoclonal Antibody (PCH101), eFluor 450, eBioscience™ |
| CD11b | BUV661 Rat Anti-CD11b Clone M1/70 |
| CD56 | Brilliant Violet 750™ anti-human CD56 (NCAM) Antibody |
| CD45RO | BB515 Mouse Anti-Human CD45RO Clone UCHL1 |
| CD25 | CD25 Monoclonal Antibody (BC96), Super Bright 600, eBioscience™ |
| PD1 | BUV737 Mouse Anti-Human CD279 (PD-1) Clone EH12.1 |
| Tim3 | CD366 (TIM3) Monoclonal Antibody (F38-2E2), Super Bright 702, eBioscience™ |
| TOX | TOX Antibody, anti-human/mouse, APC, REAfinity™ |
| TCF1 | PE anti-TCF1 (TCF7) Antibody |
| CD44 | APC/Cyanine7 anti-mouse/human CD44 Antibody |
| CD62L | BV421 Mouse Anti-Human CD62L Clone DREG-56 |
| CTLA4 | PE/Dazzle™ 594 anti-human CD152 (CTLA-4) Antibody |
| Lag-3 | CD223 (LAG-3) Monoclonal Antibody (3DS223H), PE-Cyanine5, eBioscience™ |
| Klrg1 | Brilliant Violet 510™ anti-mouse/human KLRG1 (MAFA) Antibody |
| T-bet | BV786 Mouse Anti-T-bet Clone O4-46 |
| Ki-67 | Ki-67 Monoclonal Antibody (SolA15), PerCP-eFluor 710, eBioscience™ |
| Gzmb | Granzyme B Monoclonal Antibody (N4TL33), Alexa Fluor 532, eBioscience™ |
| VISTA | VISTA Monoclonal Antibody (B7H5DS8), PE-Cyanine7, eBioscience™ |
| ICOS | Alexa Fluor® 488 anti-human/mouse/rat CD278 (ICOS) Antibody |
| CD69 | BUV805 Mouse Anti-Human CD69 Clone FN50 |

**Table S3.**
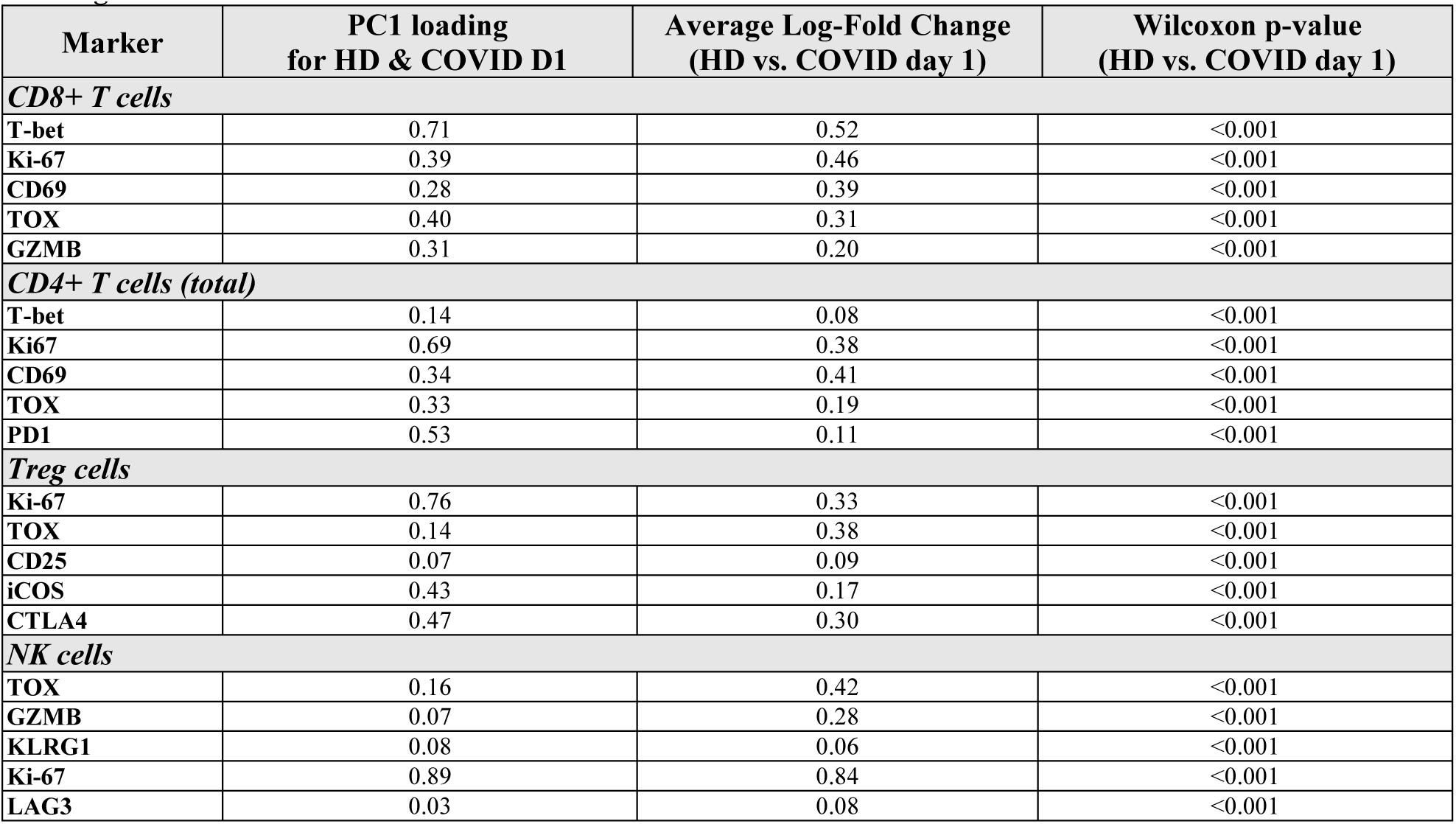
. First principal component (PC1) loadings of each activation marker were used as coefficients for defining the activation score.

| Marker | PC1 loading<br>for HD & COVID D1 | Average Log-Fold Change<br>(HD vs. COVID day 1) | Wilcoxon p-value<br>(HD vs. COVID day 1) |
| --- | --- | --- | --- |
| ***CD8+ T cells*** |  |  |  |
| T-bet | 0.71 | 0.52 | <0.001 |
| Ki-67 | 0.39 | 0.46 | <0.001 |
| CD69 | 0.28 | 0.39 | <0.001 |
| TOX | 0.40 | 0.31 | <0.001 |
| GZMB | 0.31 | 0.20 | <0.001 |
| ***CD4+ T cells (total)*** |  |  |  |
| T-bet | 0.14 | 0.08 | <0.001 |
| Ki67 | 0.69 | 0.38 | <0.001 |
| CD69 | 0.34 | 0.41 | <0.001 |
| TOX | 0.33 | 0.19 | <0.001 |
| PD1 | 0.53 | 0.11 | <0.001 |
| ***Treg cells*** |  |  |  |
| Ki-67 | 0.76 | 0.33 | <0.001 |
| TOX | 0.14 | 0.38 | <0.001 |
| CD25 | 0.07 | 0.09 | <0.001 |
| iCOS | 0.43 | 0.17 | <0.001 |
| CTLA4 | 0.47 | 0.30 | <0.001 |
| ***NK cells*** |  |  |  |
| TOX | 0.16 | 0.42 | <0.001 |
| GZMB | 0.07 | 0.28 | <0.001 |
| KLRG1 | 0.08 | 0.06 | <0.001 |
| Ki-67 | 0.89 | 0.84 | <0.001 |
| LAG3 | 0.03 | 0.08 | <0.001 |

**Table S4.**
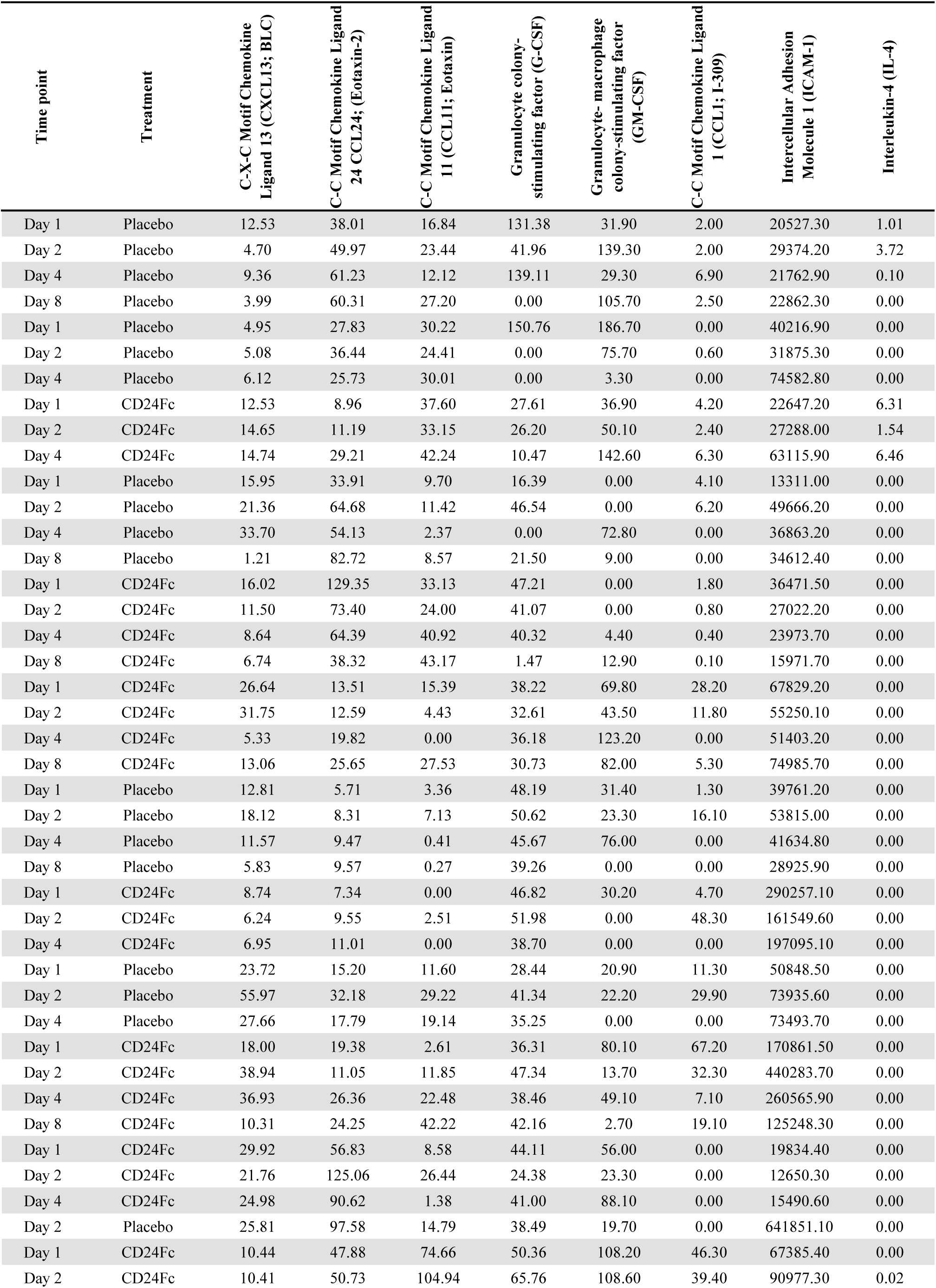
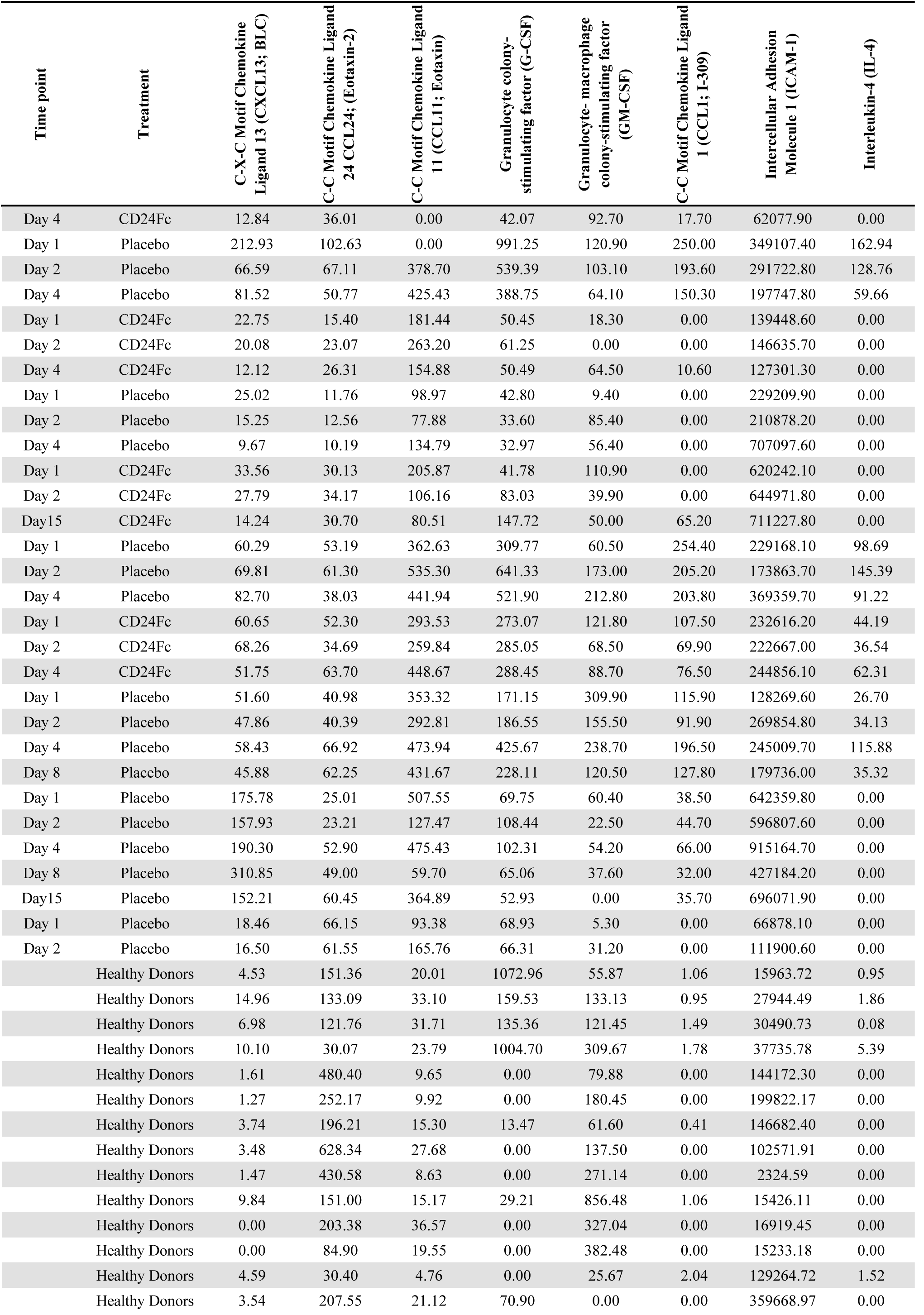
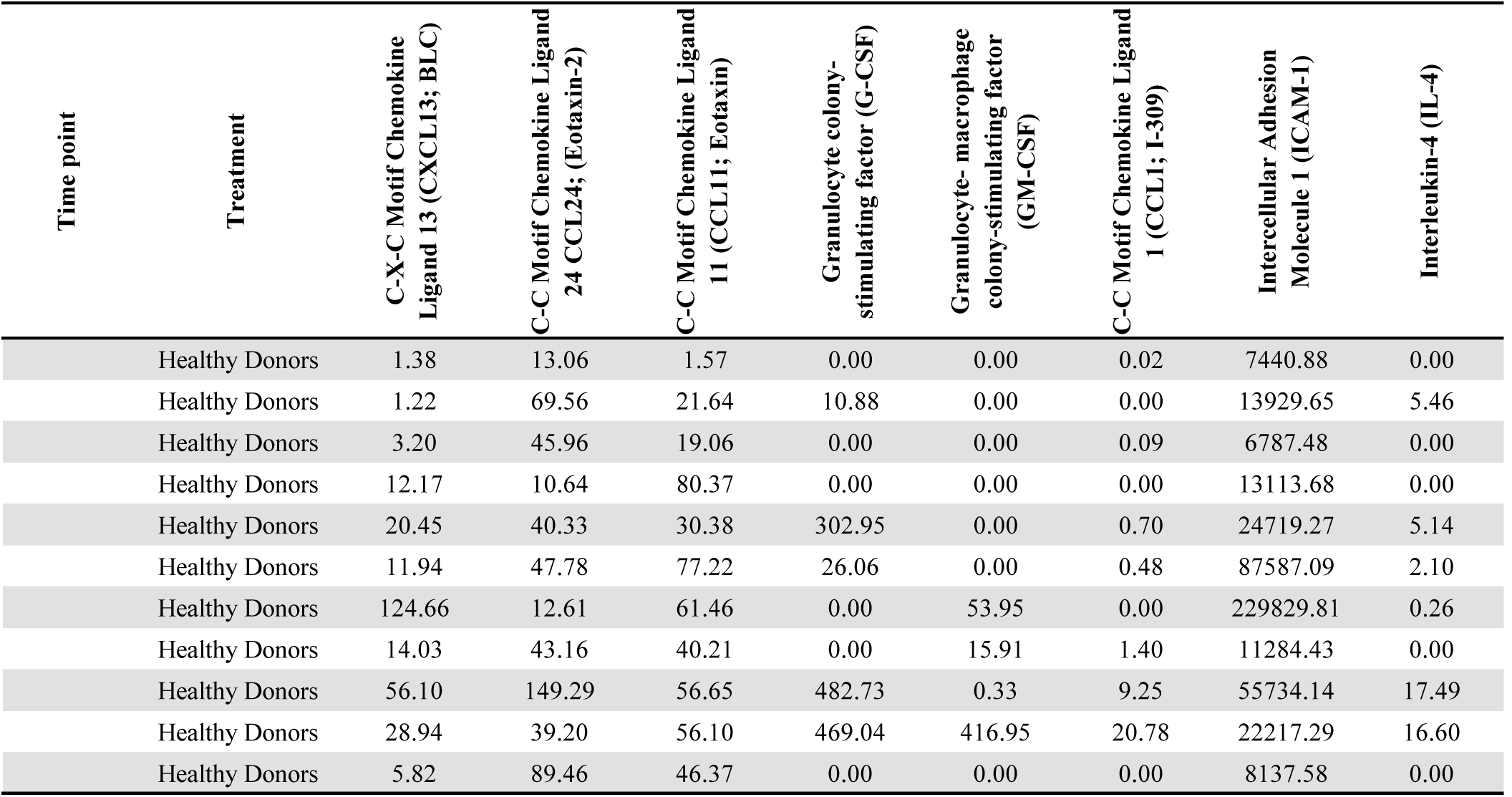
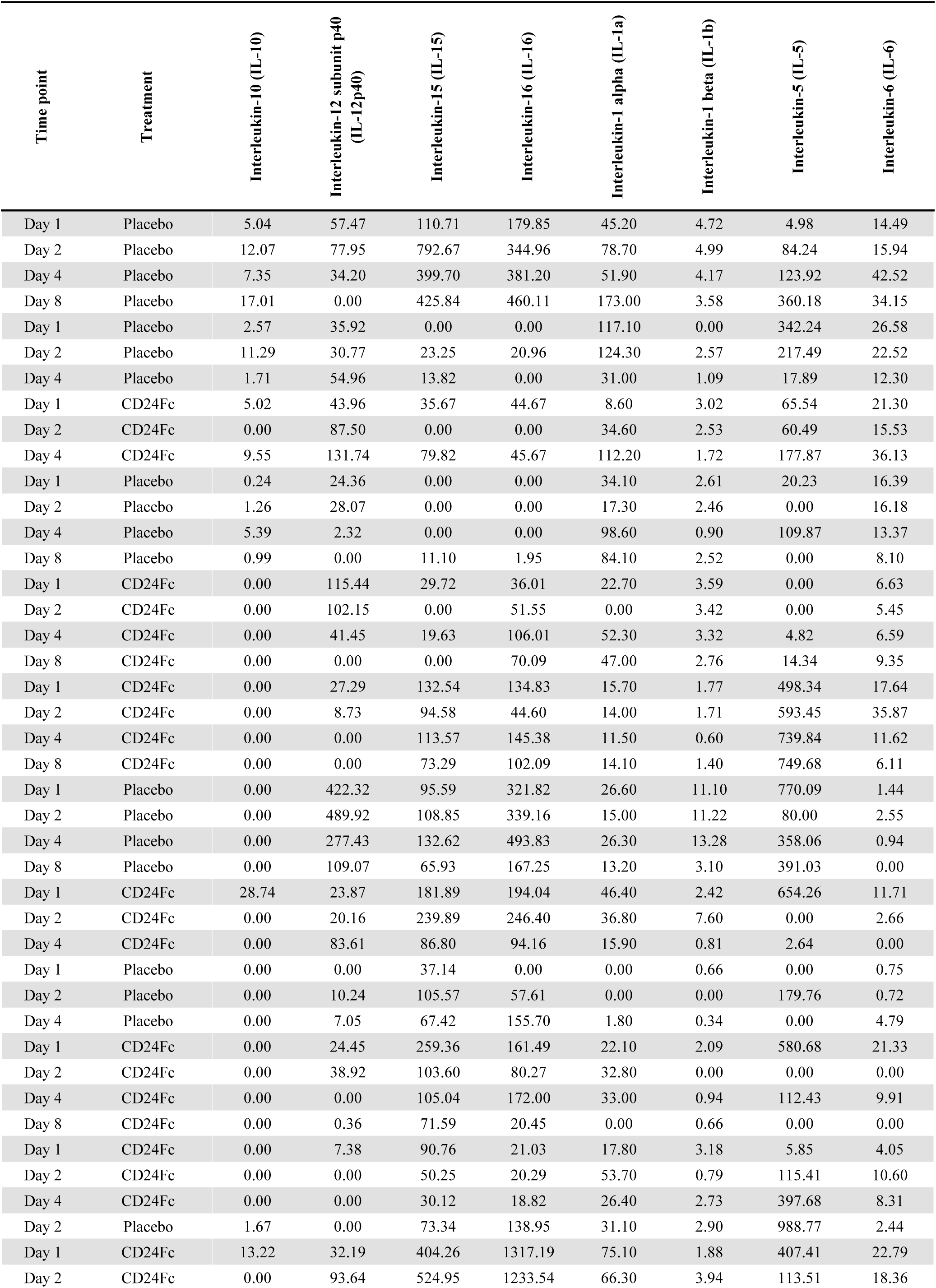
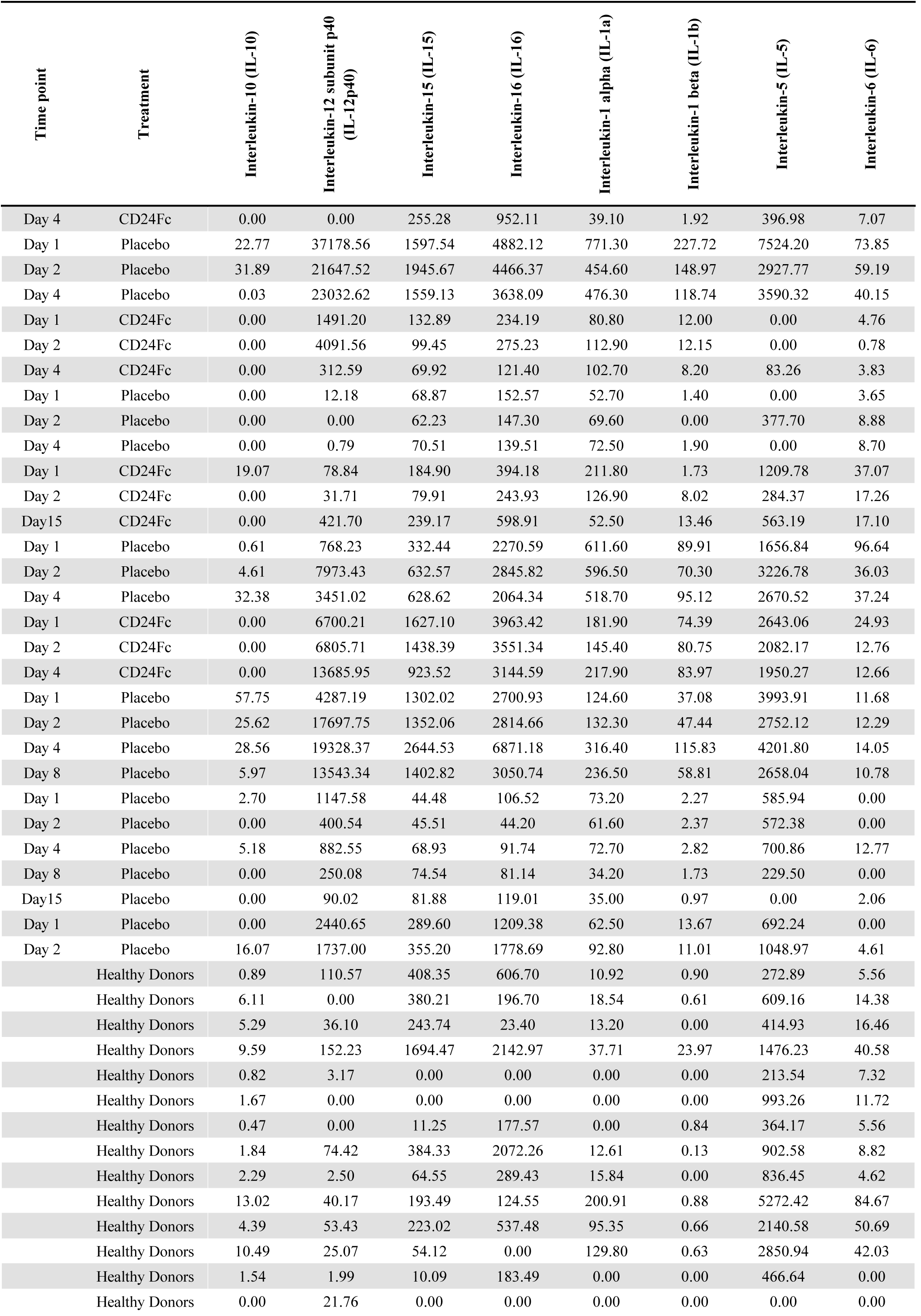
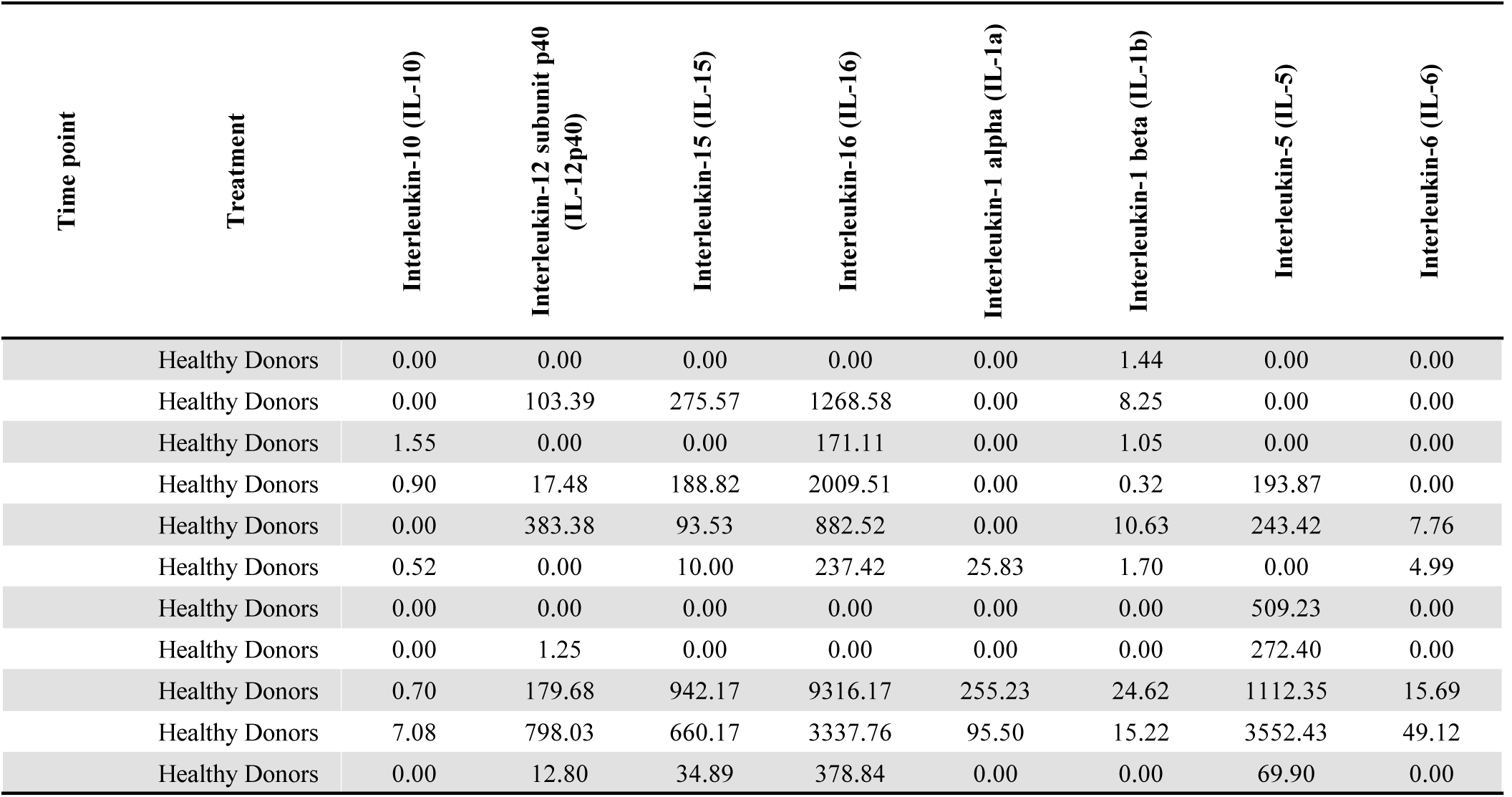
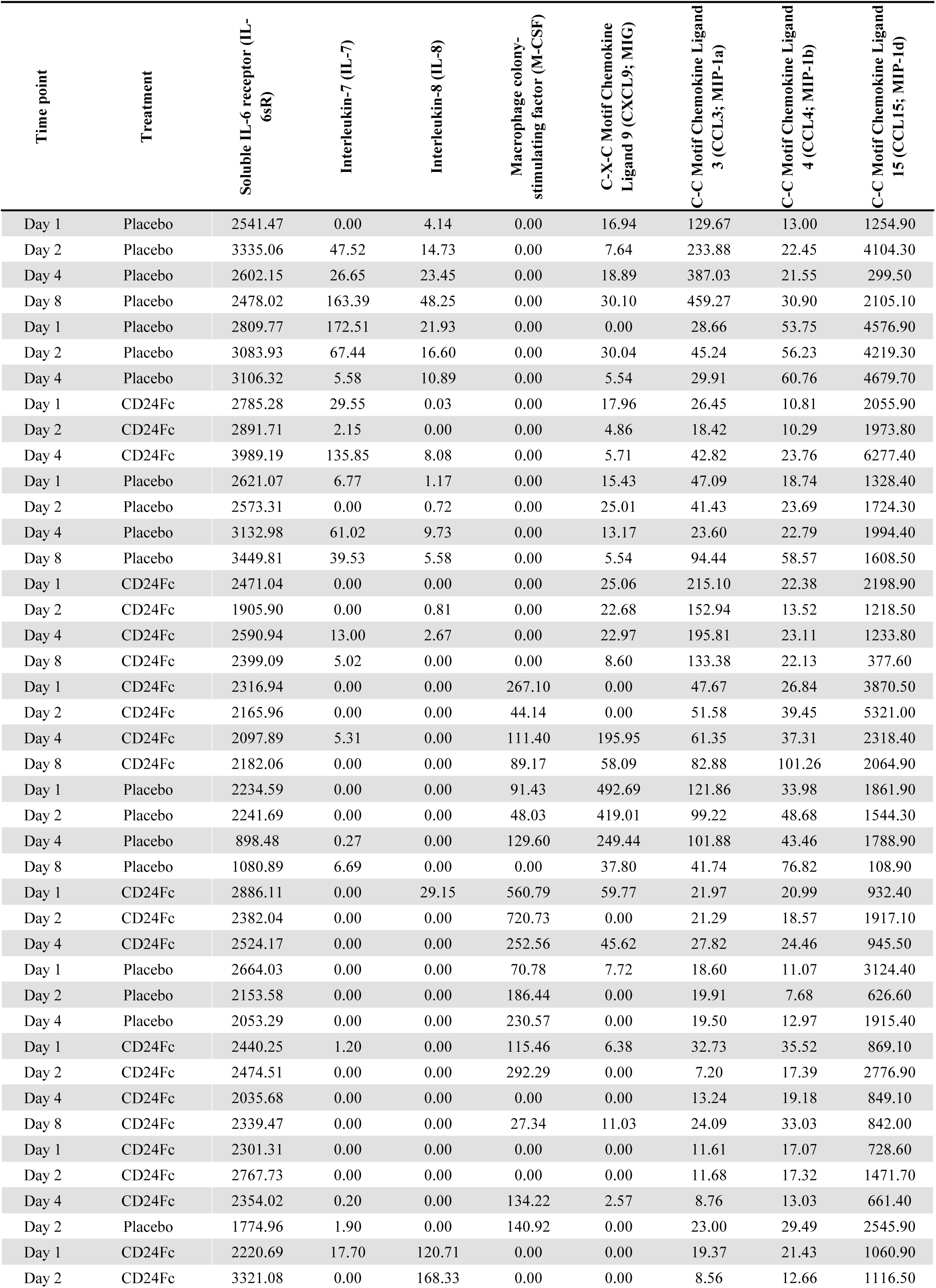
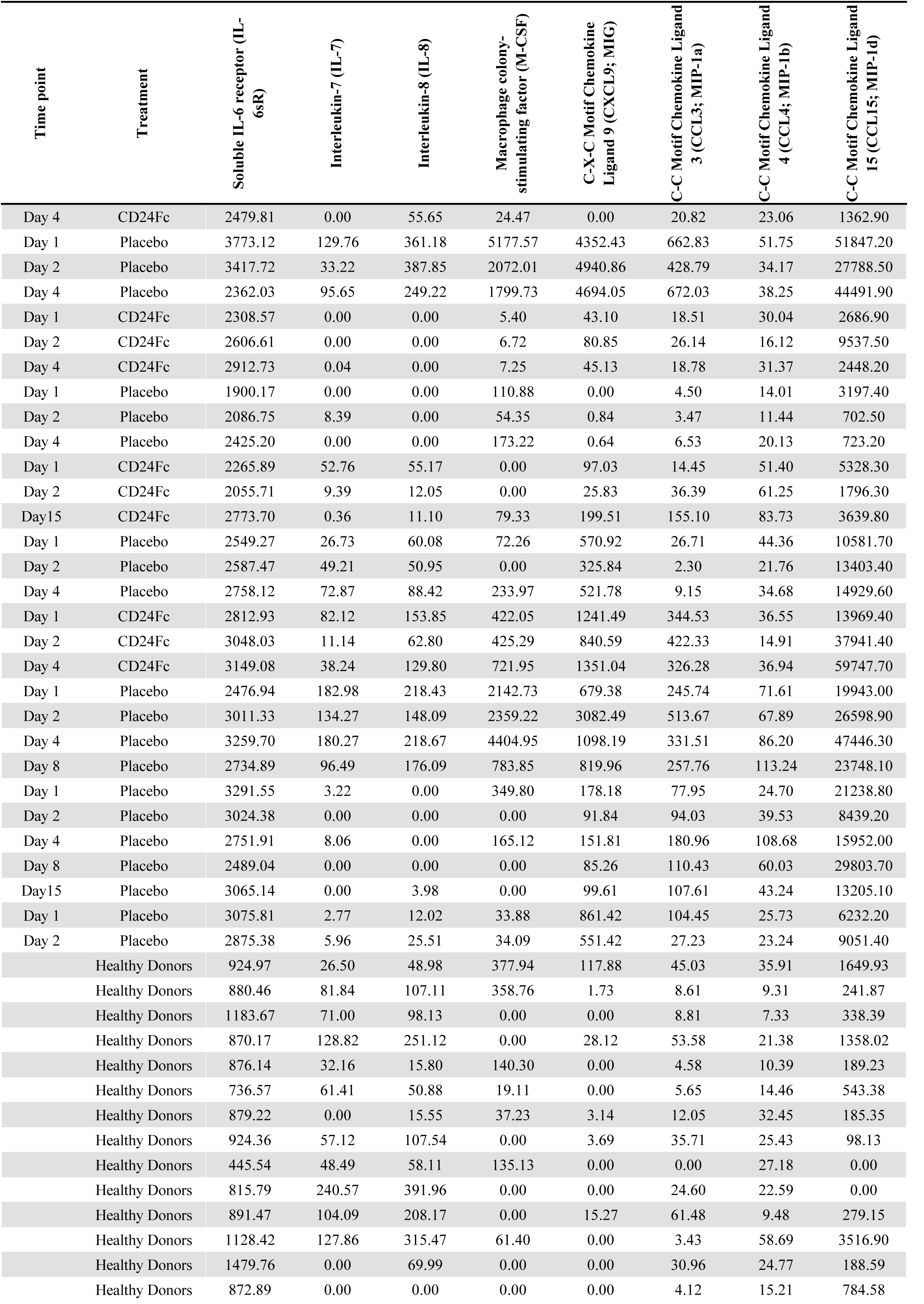
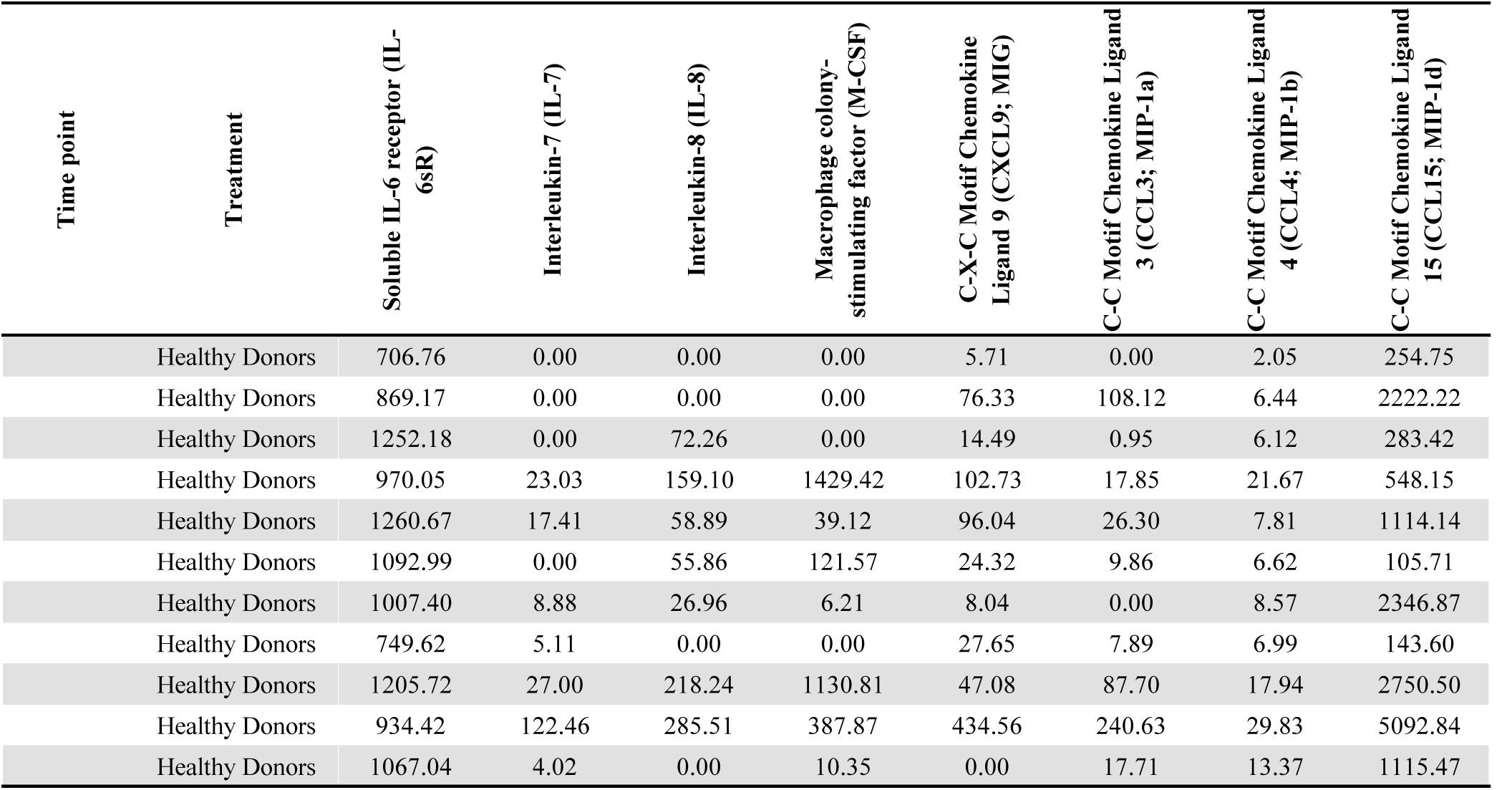
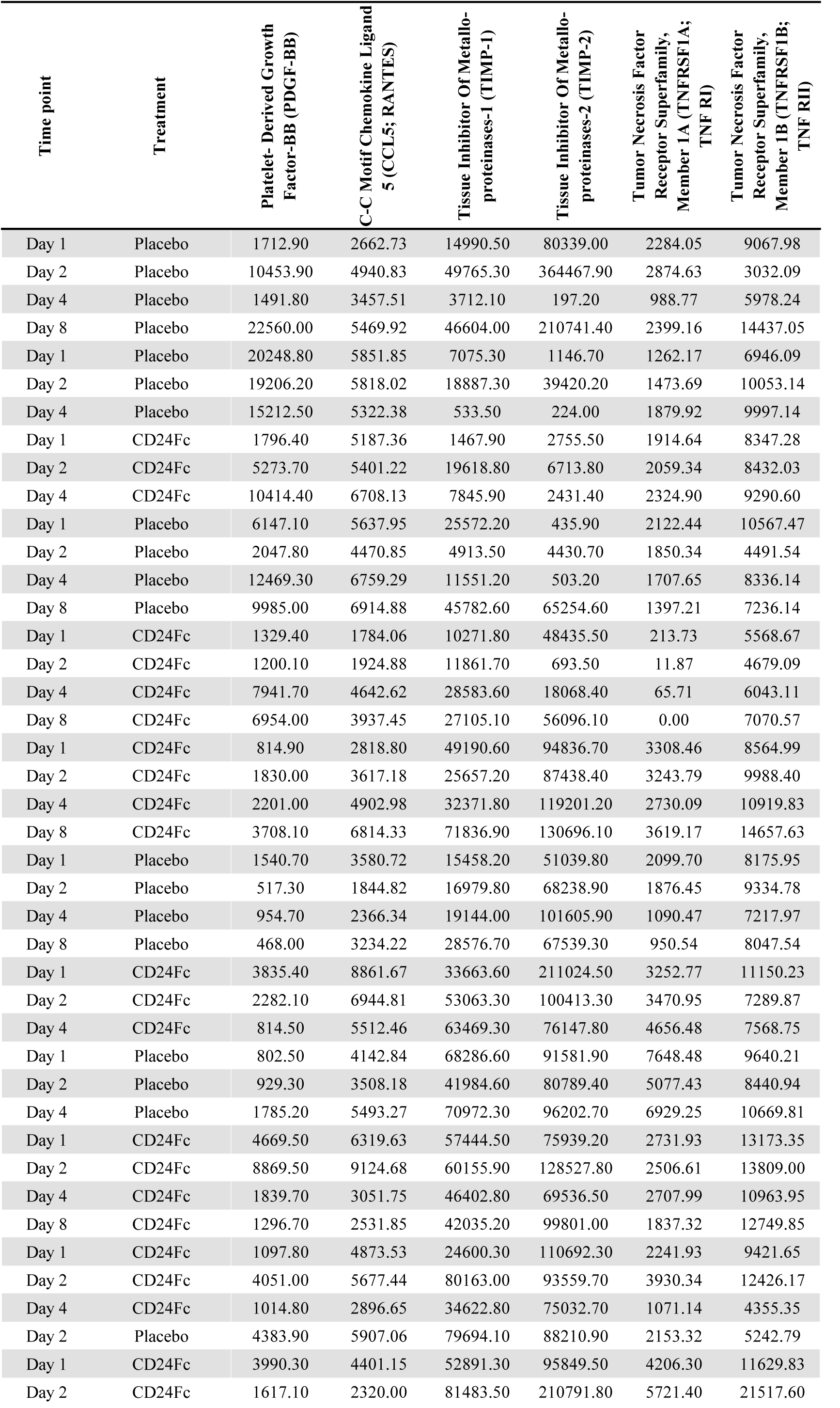
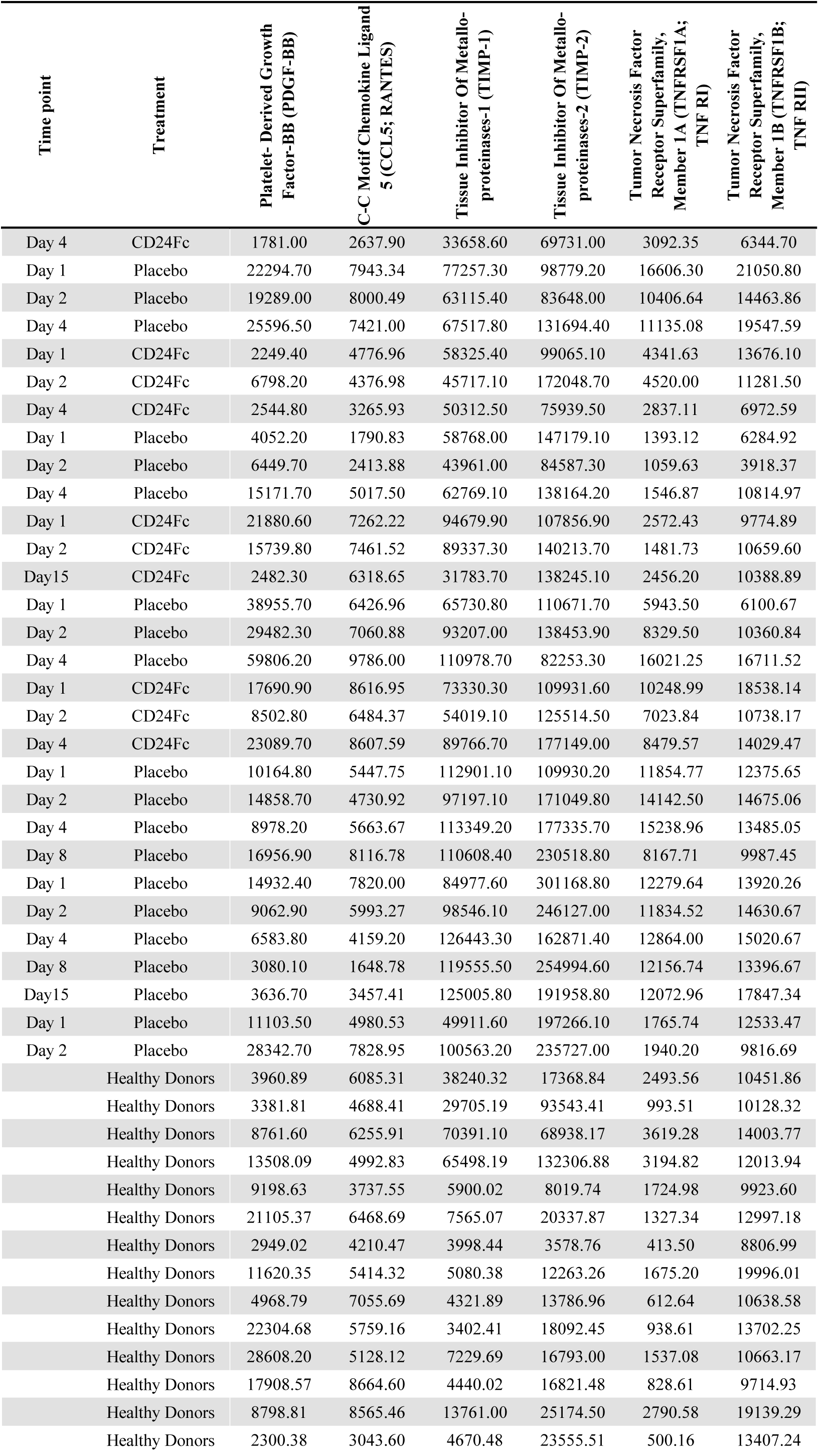
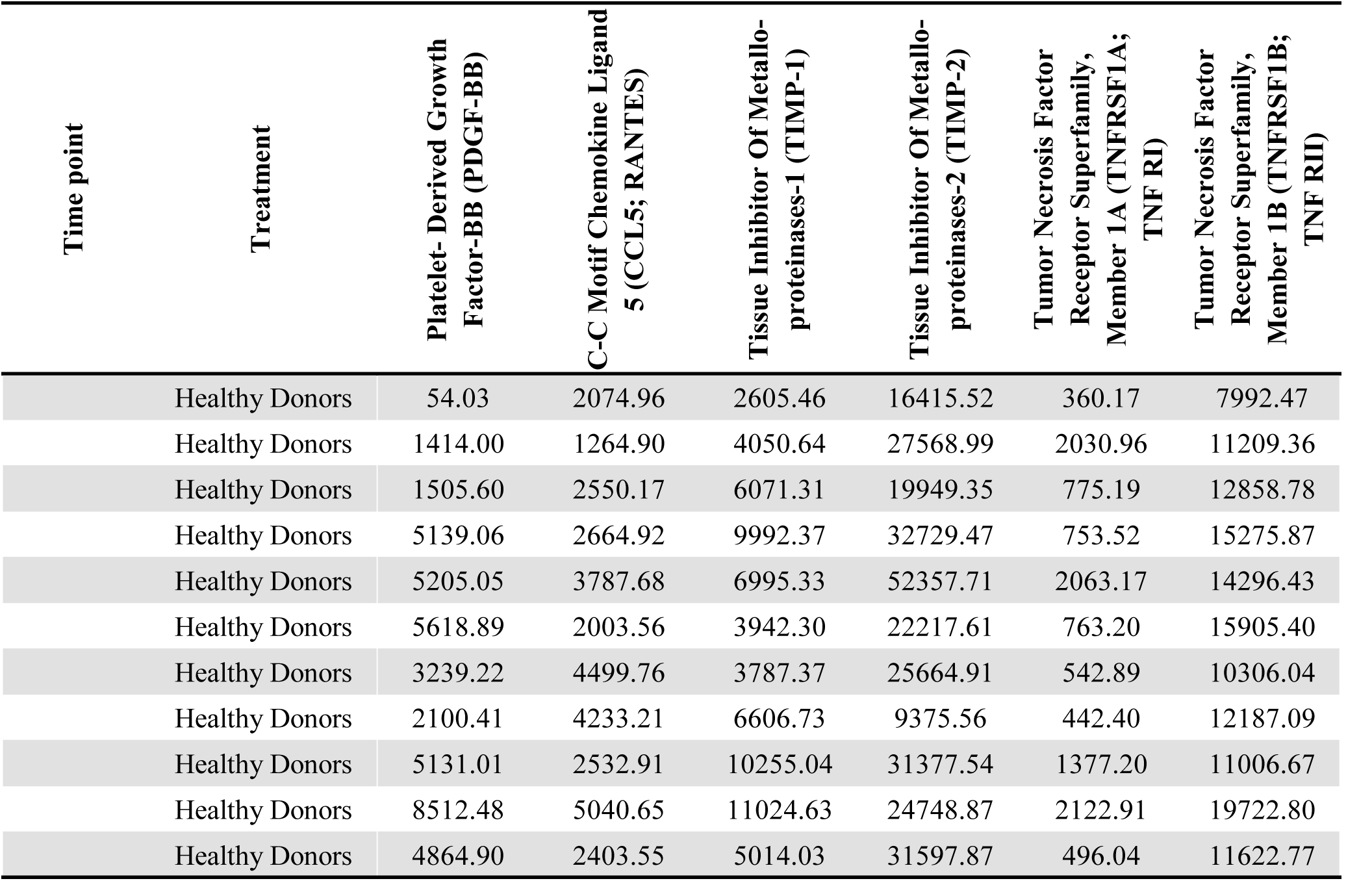
Cytokine concentrations from plasma samples (pg/mL), as measured by multiplex ELISA.

**Table S5.** Centrality ranks of filtered and weighted correlations.

| Cytokine markers | HD | D1 | Placebo | CD24Fc | var | Mean |
| --- | --- | --- | --- | --- | --- | --- |
| IL-5 | 1 | 1 | 1 | 9 | 16 | 3 |
| MIP-1d | 17 | 2 | 12 | 1 | 60·67 | 8 |
| IL-1b | 11 | 3 | 2 | 2 | 19 | 4·5 |
| IL-8 | 2 | 4 | 8 | 17 | 44·25 | 7·75 |
| G-CSF | 20 | 5 | 9 | 4 | 53·67 | 9·5 |
| IL-16 | 15 | 6 | 4 | 6 | 24·25 | 7·75 |
| MIG | 16 | 7 | 17 | 5 | 37·58 | 11·25 |
| IL-4 | 10 | 8 | 6 | 3 | 8·92 | 6·75 |
| MCSF | 24 | 9 | 13 | 16 | 40·33 | 15·5 |
| IL-12p40 | 14 | 10 | 5 | 7 | 15·33 | 9 |
| IL-15 | 13 | 11 | 7 | 8 | 7·58 | 9·75 |
| IL-1a | 7 | 12 | 11 | 12 | 5·67 | 10·5 |
| TNF RI | 25 | 13 | 14 | 14 | 32·33 | 16·5 |
| I-309 | 8 | 14 | 3 | 11 | 22 | 9 |
| MIP-1a | 12 | 15 | 21 | 13 | 16·25 | 15·25 |
| BLC (CXCL13) | 27 | 16 | 25 | 15 | 37·58 | 20·75 |
| TNF RII | 22 | 17 | 18 | 21 | 5·67 | 19·5 |
| IL-6sR | 30 | 18 | 28 | 22 | 30·33 | 24·5 |
| IL-7 | 5 | 19 | 19 | 24 | 66·92 | 16·75 |
| MIP-1b | 19 | 20 | 27 | 30 | 28·67 | 24 |
| IL-6 | 6 | 21 | 20 | 28 | 84·92 | 18·75 |
| PDGF-BB | 9 | 21 | 22 | 18 | 35 | 17·5 |
| RANTES | 18 | 23 | 23 | 19 | 6·92 | 20·75 |
| GM-CSF | 4 | 24 | 16 | 25 | 94·25 | 17·25 |
| TIMP-1 | 23 | 25 | 24 | 20 | 4·67 | 23 |
| IL-10 | 3 | 26 | 15 | 26 | 120·33 | 17·5 |
| Eotaxin-2 (CCL24) | 30 | 27 | 30 | 30 | 2·25 | 29·25 |
| ICAM-1 | 30 | 28 | 26 | 23 | 8·92 | 26·75 |
| Eotaxin (CCL11) | 26 | 29 | 10 | 10 | 103·58 | 18·75 |
| TIMP-2 | 21 | 30 | 29 | 27 | 16·25 | 26·75 |

HD, Healthy donor; D1, baseline COVID-19 patients; Var, variance.
Centrality scores ranked from highest (1, red) to lowest (30, blue). Variance and means calculated based on rank.

**Table S6.**
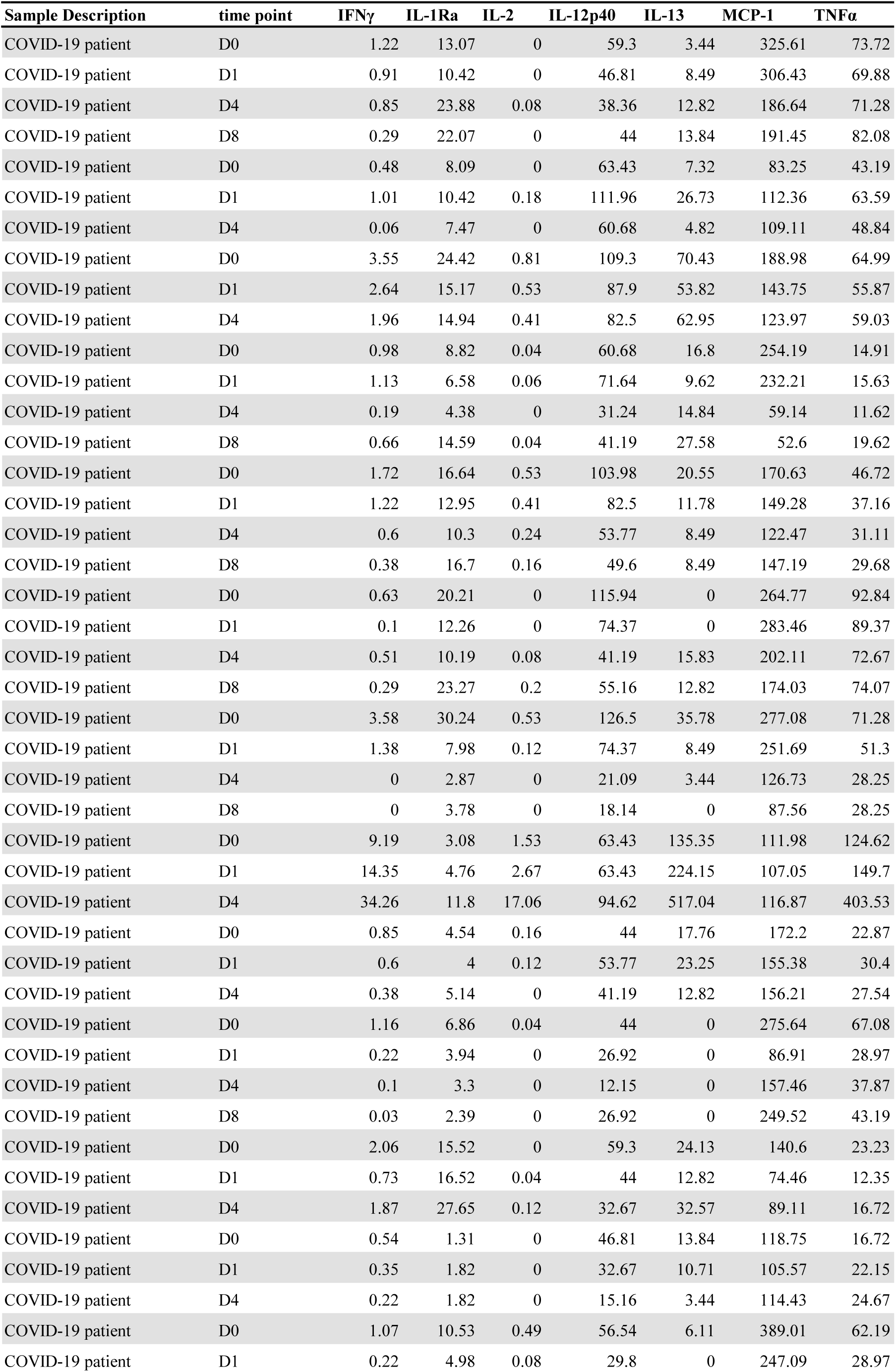
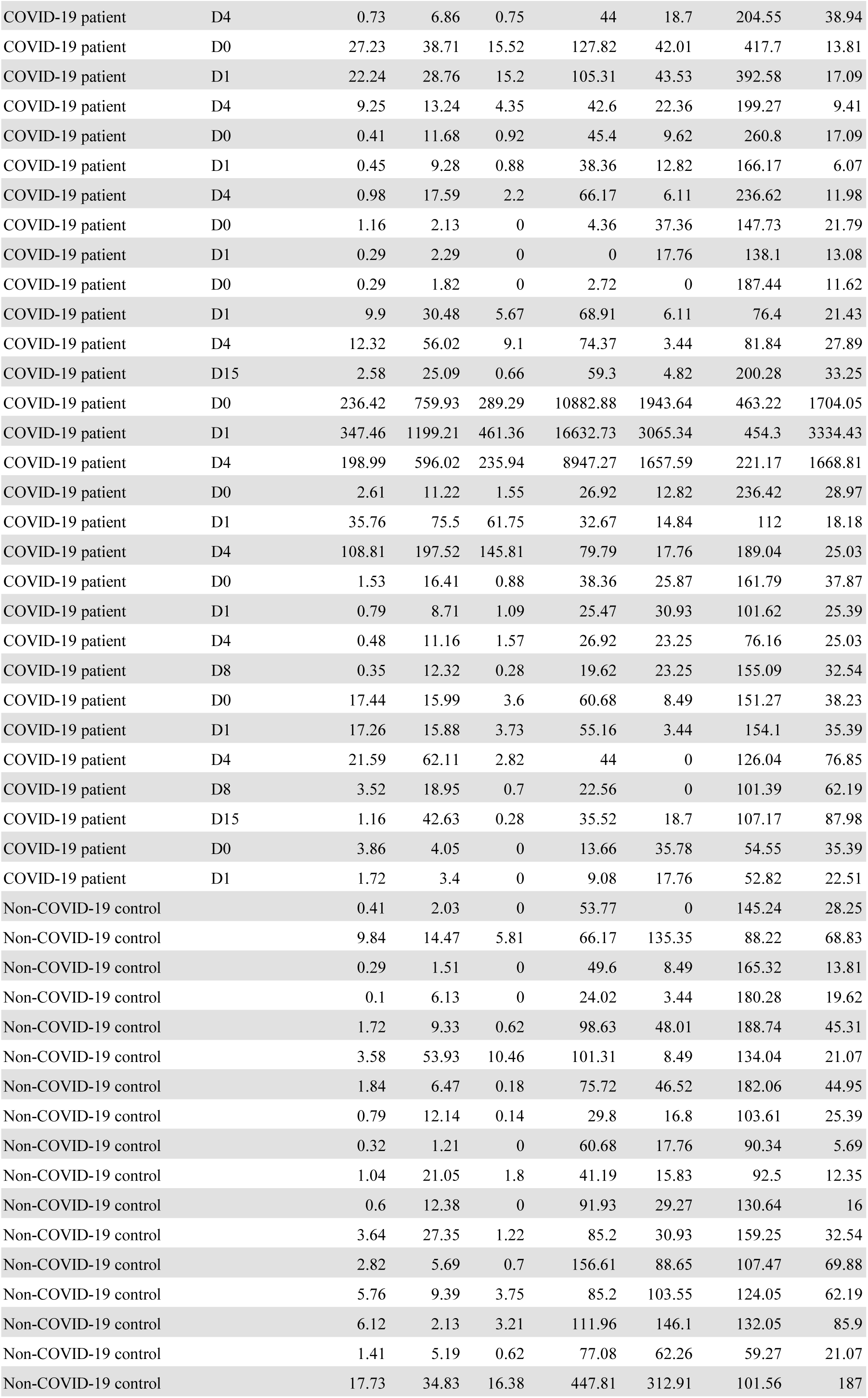
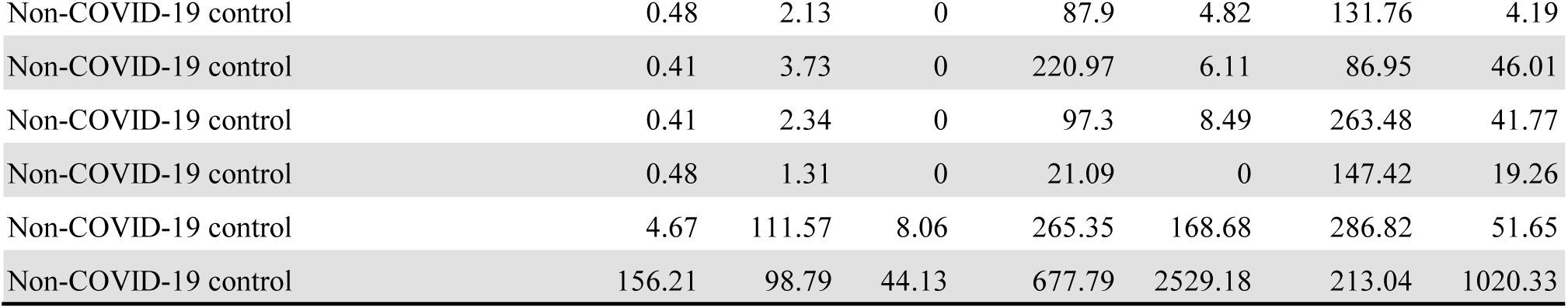
Cytokine concentrations from plasma samples (pg/mL), as measured by Luminex.

